# Determinants of SARS-CoV-2 within-host evolutionary rates in persistently infected individuals

**DOI:** 10.1101/2024.06.21.24309297

**Authors:** Mahan Ghafari, Steven A. Kemp, Matthew Hall, Joe Clarke, Luca Ferretti, Laura Thomson, Ruth Studley, Emma Rourke, COVID-19 Infection Survey Group, The COVID-19 Genomics UK (COG-UK) Consortium, Ann Sarah Walker, Tanya Golubchik, Katrina Lythgoe

## Abstract

Understanding the within-host evolutionary dynamics of SARS-CoV-2, particularly in relation to variant emergence, is crucial for public health. From a community surveillance study, we identified 576 persistent infections, more common among males and those over 60. Our findings show significant variation in evolutionary rates among individuals, driven by nonsynonymous mutations. Longer-lasting infections accumulated mutations faster, with no link to demographics, vaccination status, virus lineage, or prior infection. The nonsynonymous rate was particularly high within the N-terminal and receptor binding domains of *Spike. ORF6* was under strong purifying selection, making it a potential therapeutic target. We also identified 379 recurring mutations, with half having a negative fitness effect and very low prevalence at the between-host level, indicating some mutations are favoured during infection but disadvantageous for transmission. Our study highlights the highly heterogenous nature of within-host evolution of SARS-CoV-2 which may in turn help inform future intervention strategies.

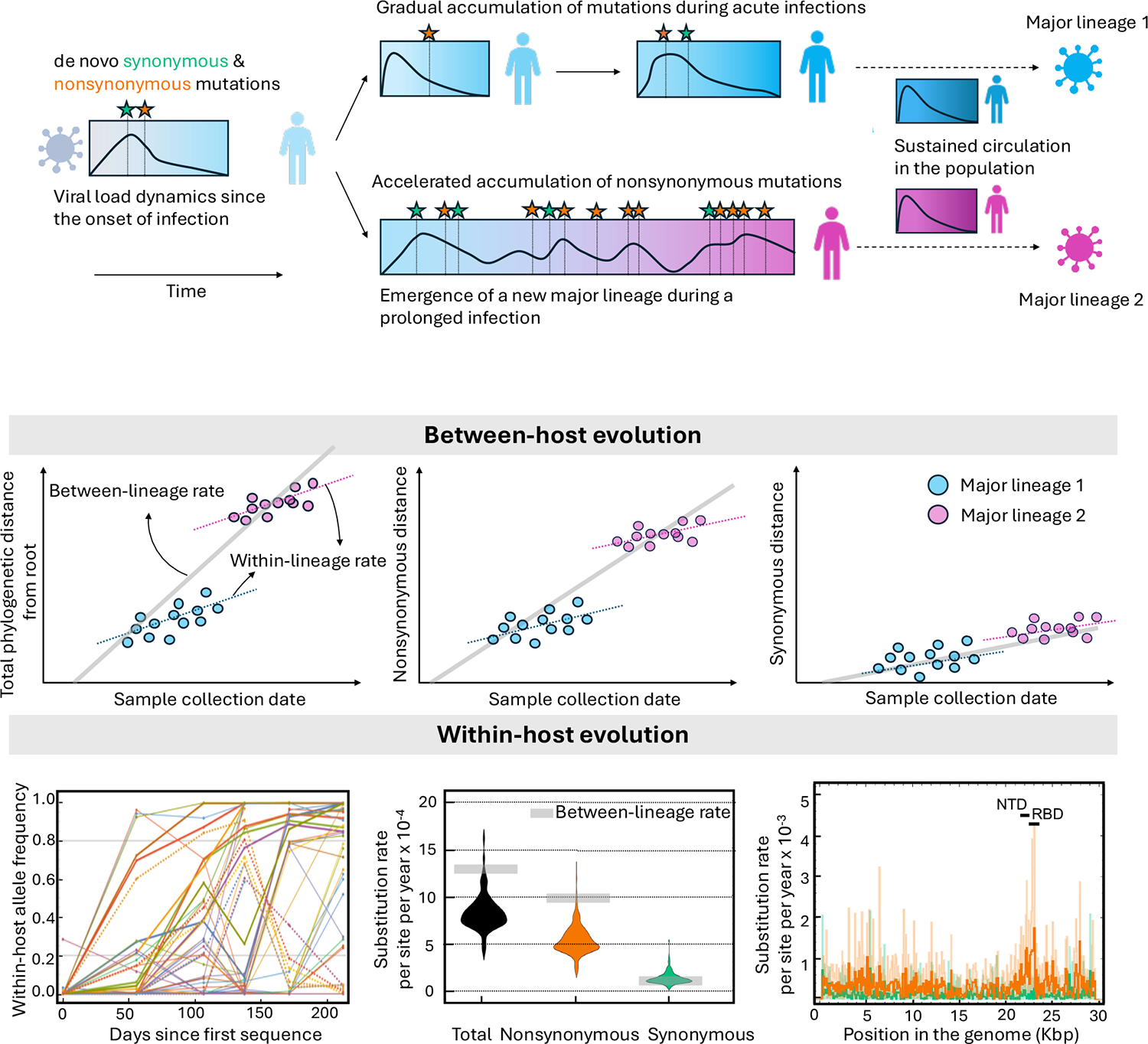

## Introduction

The evolutionary dynamics of SARS-CoV-2 has been marked by the emergence of highly divergent variants, including initial variants of concern (VOCs) Alpha, Beta, Gamma, Delta, and Omicron, followed by second-generation variants such as BA.2.75, XBB.1.5, and JN.1 ^1–3^. A notable feature of these variants is that they have a large number of nonsynonymous mutations compared to their closest ancestors, particularly in the Spike protein’s N-terminal domain (NTD) and receptor-binding domain (RBD), and show signs of strong positive selection driven by increased transmissibility and antibody immune escape ^4,5^. Within-host evolution of SARS-CoV-2 likely plays a key role in shaping these patterns of evolutionary change over time. Many chronically infected individuals also show evidence of strong viral adaptive evolution, characterised by accelerated evolutionary rates that feature key lineage-defining mutations in Spike ^1,6,7^. Given the likely importance of long-term (persistent) infections on the evolution of the virus at the population scale, we sought to characterise the evolution of SARS-CoV-2 in ‘typical’ persistent infections.

The majority of studies on the evolutionary dynamics of persistent SARS-CoV-2 infections have focussed on chronic cases. These are infections with consistently high viral titres, and are often found in hospitalised patients who are immunocompromised and receiving treatments. However, we recently showed that persistent SARS-CoV-2 infections, many of which have rebounding viral loads, are also prevalent in the general population ^8^. There remains a major gap in our understanding of host factors contributing to higher odds of experiencing persistent infections, reasons why the virus undergoes accelerated adaptive evolution in certain individuals, but not in others, identifying genomic regions and mutations, particularly outside of Spike, that undergo adaptive evolution during persistent infections, and ultimately developing effective therapeutics to clear viral infections ^9,10^. Characterisation of evolution is particularly important to determine if adaptive changes during infections mirror the saltatory evolution of SARS-CoV-2 observed with the emergence of new, highly divergent variants. In addition, identifying mutations that present complex trade-offs, being advantageous at the within-host level and detrimental at the between-host level, is crucial for understanding evolutionary factors that contribute to prolonged viral replication within hosts and increased odds of transmission between hosts ^6,11^.

Here, we explored the within-host evolutionary dynamics of SARS-CoV-2 in 576 persistently infected individuals, who participated in the Office for National Statistics Covid-19 Infection Survey (ONS-CIS), and identified factors associated with rate differences between individuals. Investigating the evolutionary dynamics of SARS-CoV-2 within persistent infections is essential for understanding the selective pressures that shape viral evolution at the within-host level, factors contributing to increased risk of resistance to treatments, and also to gauge the extent to which these individuals may contribute to the generation and subsequent spread of new variants ^12–14^.

## Results

### Saltatory evolution between major lineages for nonsynonymous but not synonymous mutations

Our analysis of the evolutionary dynamics of SARS-CoV-2 at the between-host level identifies two distinct patterns of mutation accumulation: within-lineage and between-lineage rates. Within each major viral lineage, mutations accumulate linearly over time, indicating a steady evolutionary clock (**Figure 1**; see **Methods**).

**Figure 1:**
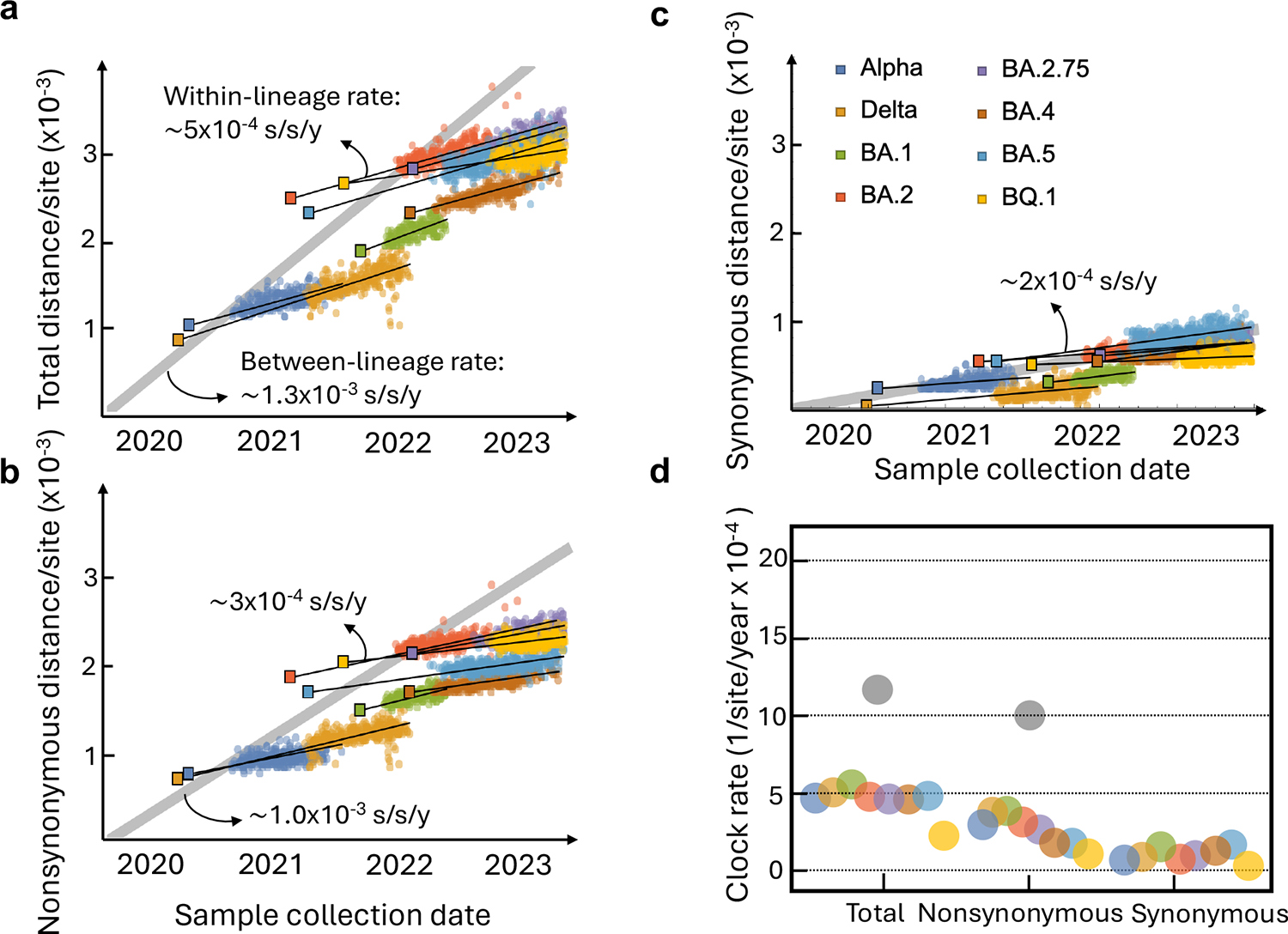
Evolutionary dynamics of SARS-CoV-2 at the between-host level. **(a)** Mutations accumulate linearly over time within each major viral lineage, punctuated by significant evolutionary leaps that demarcate these lineages (between-lineage rate; grey line). **(b)** This pattern is characterised by a disproportionate accumulation of nonsynonymous mutations at the point of transition between major lineages, whereas **(c)** synonymous mutations accumulate at a comparatively steady rate both within and across these lineages. Genetic distance within each major lineage is the Hamming distance between the putative ancestral sequence (shown with square markers) of that major lineage. The between-lineage distance is calculated as the Hamming distance between Wuhan reference sequence (NC_045512.2) and the putative ancestors of each major lineage. Lines represent the best fit from a linear regression. **(d)** Substitution rate per site per year (s/s/y) for genome-wide (total), nonsynonymous, and synonymous mutations, over time per major lineage. The substitution rates are 2.5-6.0×10^-4^ s/s/y for genome-wide, 1.5-4.0×10^-4^ s/s/y for nonsynonymous, and 0.5-2.5×10^-4^ s/s/y for synonymous mutations per major lineage. The between-lineage rate is highlighted with grey circles.

The within-lineage rate is characterised by nonsynonymous and synonymous mutations accruing at relatively similar rates. Taking synonymous mutations as a baseline for neutral changes, this suggests that the within-lineage evolution is neutral or nearly neutral. However, the evolutionary pattern is punctuated by significant leaps at the points of transition between major lineages (see Figure 1). These transitions exhibit a much higher rate of accumulation of nonsynonymous mutations compared to synonymous ones (grey line in Figure 1a-c), indicating bursts of adaptive evolution that distinguishes one major lineage from another ^15,16^.

It has long been hypothesised that this saltatory pattern of SARS-CoV-2 evolution at the between-host level comes from prolonged SARS-CoV-2 infections in immunocompromised individuals, where the virus has extended time to adapt and accumulate advantageous mutations without undergoing tight transmission bottlenecks, followed by the onward transmission of the highly divergent virus to the rest of the population ^1^. We set out to investigate whether viral evolution in long infections is consistent with this hypothesis by analysing the evolutionary dynamics of SARS-CoV-2 in 576 persistently infected individuals identified as part of the ONS-CIS.

### Persistent infections are more frequent in older individuals and males

We defined persistent infections as those with at least two RT-PCR positive samples with a high viral RNA titre (cycle threshold values ≤30), collected at time intervals of at least 26 days apart, and representing the same infection. We previously identified 381 persistent infections within the ONS-CIS using samples collected between 2 Nov 2020 to 15 August 2022 ^8^. For the current analysis, we extended this dataset to samples collected up to 21 March 2023, and so covering the entire duration of ONS-CIS before it was paused ^2^, and thereby identifying an additional 195 persistent infections (see **Methods**).

In total, our dataset comprised 576 cases of persistent SARS-CoV-2 infections, including 11 infections with B.1.1.7 (referred to as Alpha), 106 B.1.617.2 (referred to as Delta), 102 with BA.1, 204 with BA.2,16 with BA.4, 133 with BA.5, and 4 with XBB major lineages. All persistent infections had viral sequencing data from at least two time points; 27 had sequencing data from three or more time points, typically collected at 20-to 40-day intervals, and the longest-lasting persistent infection spanned nearly a year with eight sequenced time points (Figure 2a-c).

**Figure 2:**
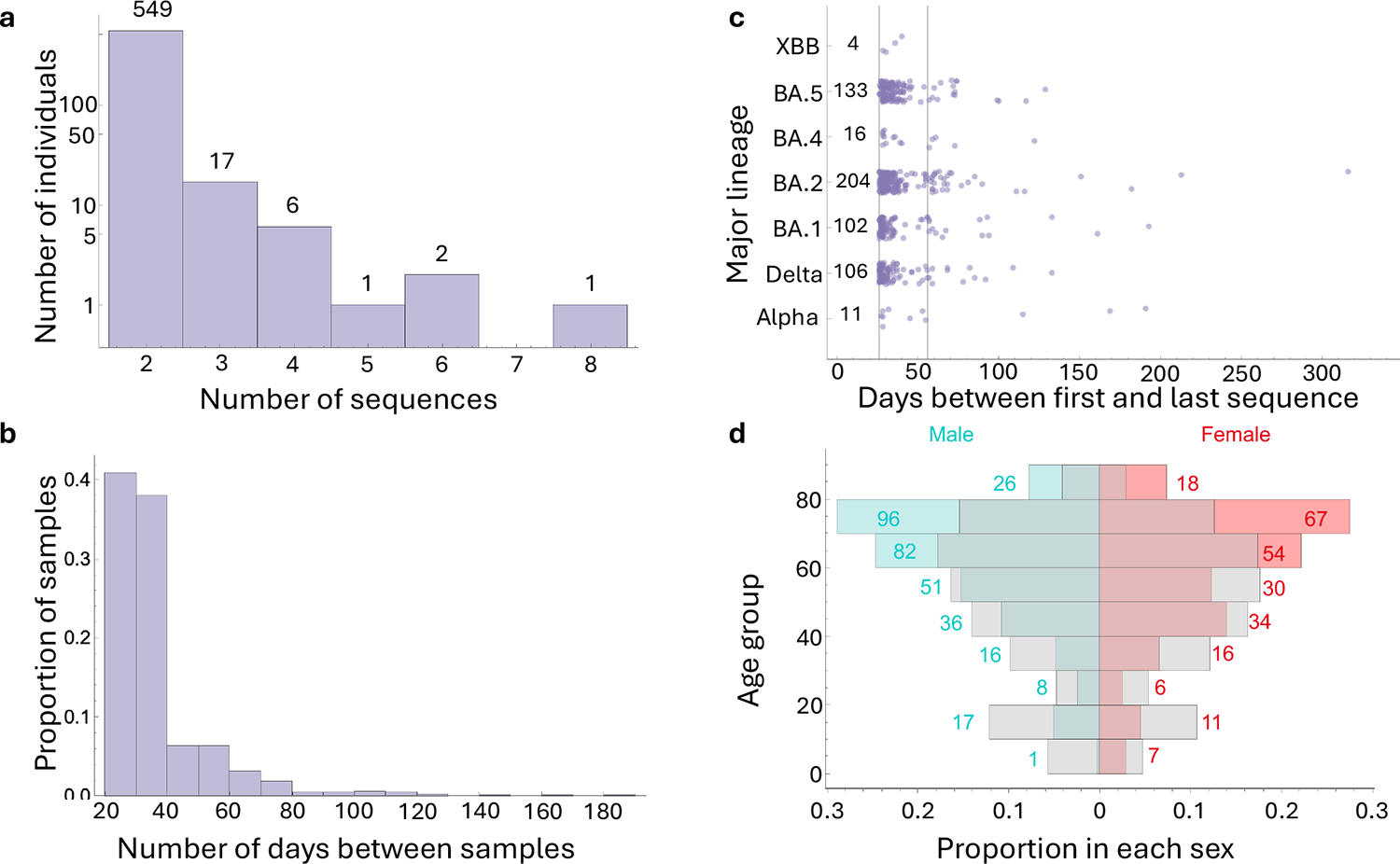
Baseline characteristics of persistent SARS-CoV-2 infections. **(a)** Number of sequences per persistent infection. Numbers on each bar show the number of persistent infections per category. **(b)** Distribution of numbers of elapsed days between consecutive sequences collected per persistent infection. In cases where a persistent infection has multiple samples, each pair of consecutive samples is considered. **(c)** Number of days between the earliest and latest genomic samples for each persistent infection, with each point representing a persistent infection. Solid vertical lines are drawn at the 26- and 56-day marks to denote the thresholds for persistent infections lasting at least one month and two months, respectively.

Numbers on the side of each bar shows the total number of persistent infections per major lineage. **(d)** Proportion of persistent infections in each sex and per age-group. Numbers on each bar show the raw number of persistent infections in each age-group. Grey bars on either side show the relative proportion of infections with a single positive PCR within the ONS COVID Infection Survey per sex and age group.

Compared to individuals with a single positive PCR test within the ONS-CIS (hereafter referred to as non-persistent infections), persistently infected individuals were more prevalent in the above 60 age groups (X-squared = 8.98, df = 1, p-value = 0.00273; see also Figure 2d). We also found a significant association between sex and type of infection (X-squared = 21.28, df = 1, p-value = 3.97×10^-6^), with males representing 57.8% of persistently infected cases compared to 48.1% of non-persistently infected cases. Although we lacked specific information about the underlying health conditions of participants, the age and sex profile of individuals with persistent infections closely mirrors the demographic characteristics of individuals diagnosed with Type 1 and Type 2 diabetes in England ^17^.

### Nucleotide diversity increases during infection

We began investigating the within-host evolutionary dynamics of the virus in these 576 individuals by first identifying intra-host single nucleotide variants (iSNVs) for each sample collected during infection, and measuring nucleotide diversity, *π*, over time (see **Methods**). An iSNV was called at a given genomic position if there was a minimum read depth of 10 at that position and a minor allele present at frequency of 20-50%. Positions where the majority of reads were gaps, and those where observed iSNVs are unlikely to represent genuine within-host diversity, were excluded from the analysis (see **Methods**).

In the great majority of cases, nucleotide diversity at the earliest time point for each persistent infection was very low, with more than 61% (355/576) of infections displaying no detectable diversity (Figure 3a). This suggests that the first sample in most persistent infections was collected near the onset of infection, and with infection initiated by a single, or very closely related, variants ^18,19^. The average within-host diversity (*π*) of all sampling time points was approximately 4×10^-5^ per nucleotide which is more than an order of magnitude smaller than the between-host diversity at approximately 5×10^-4^ per nucleotide ^2^. As might be expected, this indicates that samples collected from the same infection have much lower diversity than samples collected independently from different individuals ^20^. Despite significant variation in diversity over time across different infections (Figure 3b), genetic diversity tended to increase until approximately 100 days after the first time point, at which point it either declined or began to plateau in most cases. This pattern suggests that iSNVs appearing late in the infection do not significantly contribute to the overall nucleotide diversity. This could be because they reach mutation-selection balance, remain at low frequency due to their deleterious fitness effects, or rapidly increase in frequency and become fixed. A similar pattern has also been observed during the within-host evolution of HIV ^21^.

**Figure 3:**
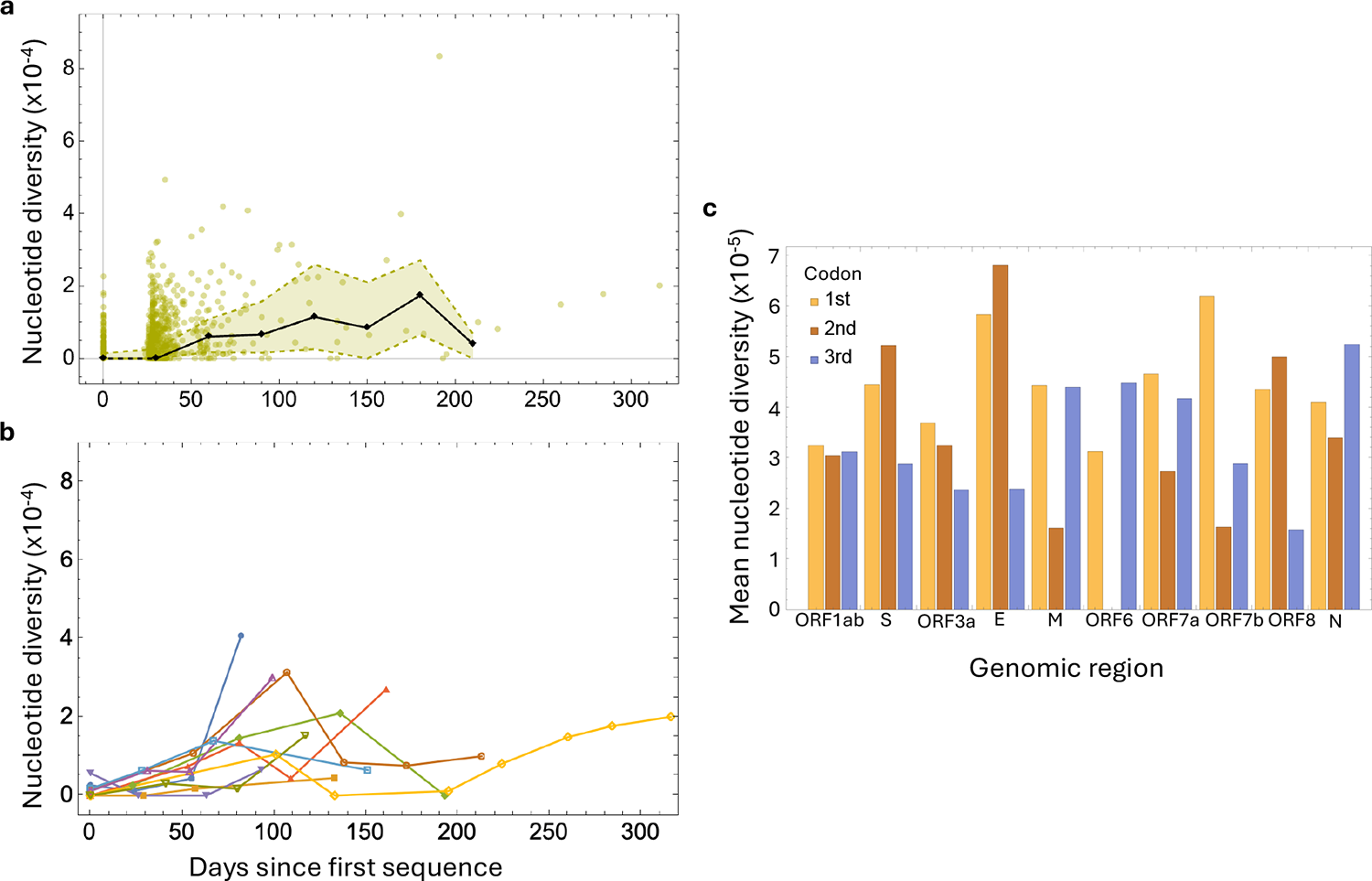
Within-host nucleotide diversity. **(a)** Aggregate nucleotide diversity (*π*) over time across all persistent infections. Each data point represents the diversity of a sample from a persistent infection at a given time since the first sequenced sample in that infection (*t=*0). The black line shows the median nucleotide diversity in 30-day intervals and the shaded area covers the interquartile range. **(b)** Nucleotide diversity over time for persistent infections with three or more samples. **(c)** Mean nucleotide diversity per codon position in each genomic region including the Open Reading Frames (ORFs), *Spike* (*S*), *Envelope* (*E*), *Membrane* (*M*), and *Nucleocapsid* (*N*).

We also measured nucleotide diversity by codon position. The first and second codon positions typically induce nonsynonymous changes, while most mutations in third position result in synonymous changes ^22^. Looking at the first and second position across different genomic regions within our samples from persistent infections, the lowest nucleotide diversity was in open reading frame 6 (*ORF6*), with no diversity at the second position, indicating this genomic region is highly conserved and likely subject to strong purifying selection. Conversely, the *Envelope* (*E*) gene exhibited the highest diversity at the first two codon positions, followed by *Spike* (*S*) and *ORF8* (Figure 3c). Some of the other genomic regions such as *ORF1ab* had a more uniform diversity across all three codon positions while *ORF6* and *Nucleocapsid* (*N*) had higher synonymous diversity compared to nonsynonymous diversity across all genomic regions.

### Higher prevalence of nonsynonymous mutations later in infection

Next, we identified synonymous and nonsynonymous mutations present at 20% frequency or above at any time point over the course of infection, taking the majority allele at the first time point as reference (see **Methods**). Nearly 67% of all mutant alleles and 73% of those within the coding region were nonsynonymous, with less than 2% synonymous at the first and second codon positions (Figure 4a). *ORF6*, *Membrane* (*M*), and *N* had the highest proportion of synonymous compared to nonsynonymous mutations, and *ORF8* the lowest (Figure 4b). Comparing the allele frequency of mutations at different points during infections, towards the start of infections (less than 120 days since the first sampled time point), both nonsynonymous and synonymous alleles were typically at comparable frequencies, predominantly below 50% (Figures 4c,**d**). However, later on nonsynonymous alleles tended to be at higher frequencies, likely indicative of positive selection (see Figure 4c**,d**).

**Figure 4:**
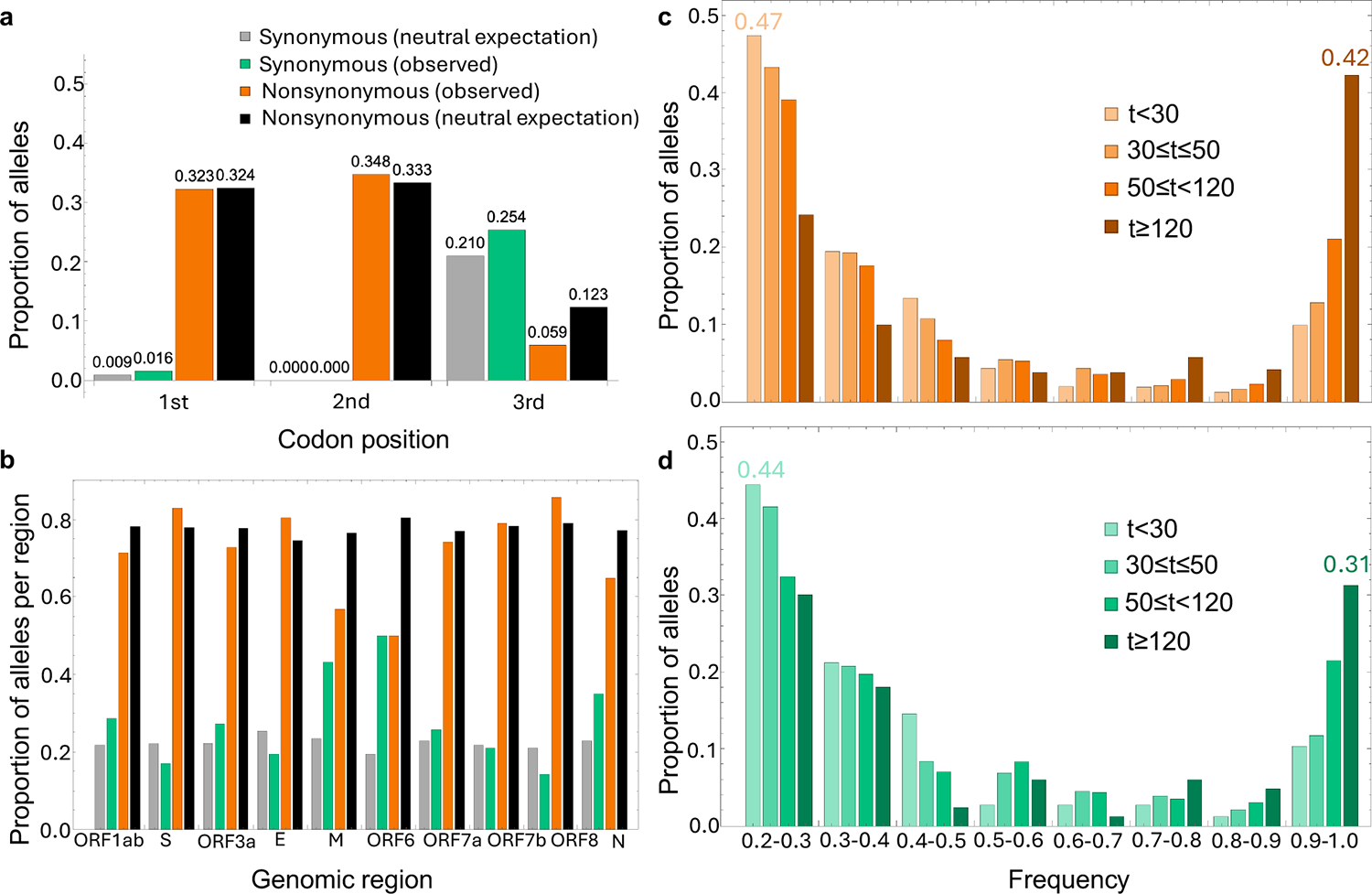
Basic characteristics of mutant alleles. **(a)** Proportion of synonymous (green) and nonsynonymous (orange) mutant alleles per codon position observed in samples from persistent infections, taking the majority allele at the first time point as reference, compared to expectations under neutrality, taking NC_045512.2 as reference. **(b)** Proportion of alleles per mutation type for each genomic region including the Open Reading Frames (ORFs), *Spike* (*S*), *Envelope* (*E*), *Membrane* (*M*), and *Nucleocapsid* (*N*). **(c)** Proportion of synonymous and (**d**) nonsynonymous alleles over time across different frequency bands. The proportions of alleles within the smallest and largest frequency bands are highlighted for both early (t<30) and late (t≥120) stages of infection.

Nonsynonymous alleles were two to three times more prevalent than synonymous ones across all frequency bands (see Supplementary Figure 1), with about 73% of mutants in the coding region that exceeded 50% frequency being nonsynonymous. This ratio is close to the expectation under neutrality, with 78% of all possible mutations across the genome expected to be nonsynonymous ^22^ (see Figure 4a**,b**). Given it has previously been found that half of the mutations causing nonsynonymous changes are purged both at the between-host level and during acute infections (dN/dS ≈0.5) ^18,23^, observing a ratio of nonsynonymous mutations that is similar to the neutral expectation in persistently infected individuals suggests that at least some genomic regions are under positive selection.

### Variation in evolutionary rates among infections is driven by nonsynonymous changes

To determine the within-host evolutionary rates for each infection, we used changes in allele frequency relative to first sequenced time point (hereafter referred to as the baseline) as a proxy for measuring evolutionary distance over time (see **Methods**). Within this framework, a full sweep of a mutant allele (a frequency change of 100%) contributes 1 unit of distance and a partial sweep with a frequency change of 40% contributes 0.4 units. This definition of evolutionary distance does not invoke any assumptions about the founder population, which might differ from the population at baseline, as it relies on absolute changes in allele frequencies to measure evolutionary distance ^24^.

Allele frequencies change over the course of infection both as a result of sampling noise and actual evolution. To assess the impact of sampling noise on the variation in allele frequencies over time, we required that at least one allele be present at a frequency of ≥20% at at least one time point per persistent infection. In other words, if no allele meets this threshold in any sample from a persistently infected individual, we will not (incorrectly) assume there is zero noise (or evolution) due to insufficient data. We also limited our analysis to samples with sufficient sequencing coverage to ensure unbiased estimates of genetic distance per site and, therefore, excluded samples where the number of overlapping base pairs between the consensus sequence at the baseline and the consensus sequence of the sample was less than half the length of the genome. Approximately 14% (82/576) of persistent infections did not meet these criteria and were excluded. We categorised the genetic distances as either synonymous or nonsynonymous, depending on whether the mutant alleles induced a synonymous or nonsynonymous change to the consensus sequence at the baseline for each persistent infection.

To determine within-host evolutionary rates we used linear regression models, with the slope of the regression line representing the rate of evolution and the y-intercept the level of background noise in the data. The non-zero y-intercept could be attributed to sampling noise and/or residual population structure at baseline ^24^. To determine the most appropriate model for measuring within-host evolutionary rates, we compared several linear regression models with varying levels of complexity based on their Bayesian Information Criterion (BIC) values. This comparison included a null model which assumed a single fixed slope and y-intercept for all persistent infections (see **Methods**).

For genome-wide and nonsynonymous genetic distances, a linear mixed-effect model which assigned a unique evolutionary rate to each persistent infection, but a fixed y-intercept for all infections, gave the best fit (**Supplementary Table 1**). For synonymous distances, a model with a single rate for all infections, but a random y-intercept for each infection, was the most appropriate model. This suggests there was considerable variation in the rate of evolution among individuals, predominantly influenced by nonsynonymous changes, and no strong evidence supporting variation in rate of synonymous evolution across individuals. We also confirmed that the level of noise in allele frequencies is not associated with different sequencing centres (see **Methods** and **Supplementary Table 1**).

The median genome-wide evolutionary rate was 7.9×10^-4^ substitutions per site per year (s/s/y) with an interquartile range (IQR) of 7.0-9.0×10^-4^ s/s/y (Figure 5a). Almost 95% (469/494) of persistent infections exhibited an evolutionary rate exceeding 5.5×10^-4^ s/s/y, indicating that the vast majority of individuals experienced a rate surpassing the between-host within-lineage evolutionary rate of SARS-CoV-2 which typically ranges from 2.5 to 5.0×10^-4^ s/s/y for the Alpha, Delta, and Omicron sublineages (see Figure 1). Furthermore, 23% (114/494) of the infections had an evolutionary rate higher than the between-lineage rate of 1×10^-3^ s/s/y. The rate of nonsynonymous evolution was 5.0×10^-4^ (IQR: 4.4-6.1×10^-4^) s/s/y, which was about four times higher than the synonymous rate of 1.2×10^-4^ s/s/y across most persistent infections (see Figure 5).

**Figure 5:**
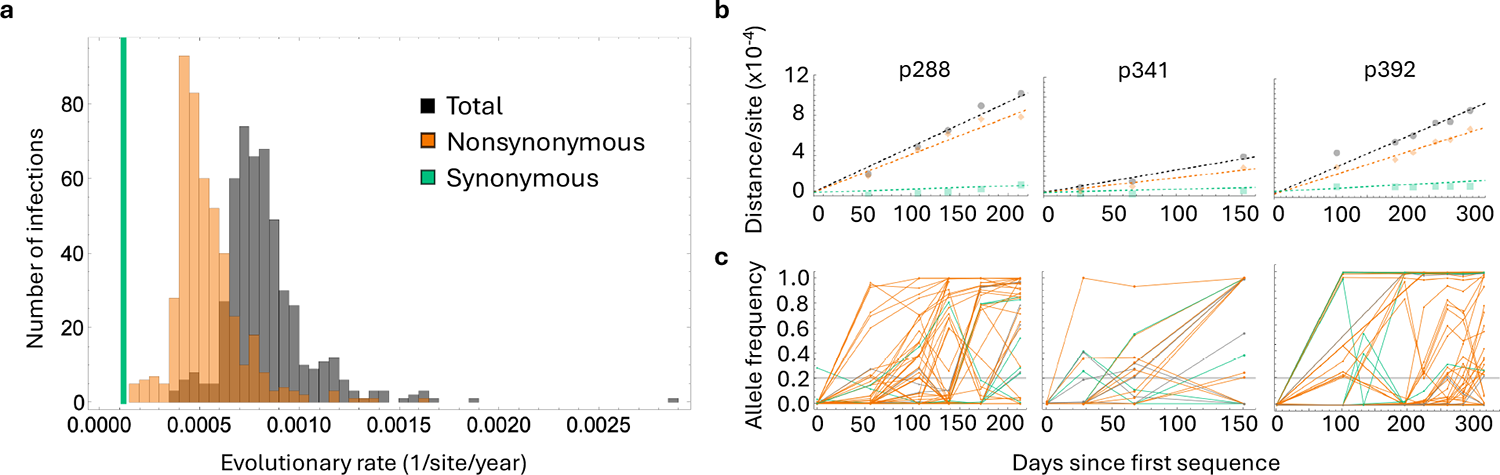
Rates of genome-wide, nonsynonymous, and synonymous evolution in persistently infected individuals. **(a)** Distribution of inferred evolutionary rates per individual, based on analyses using a linear mixed-effects model optimised for the best fit to the data (as indicated by the lowest BIC value). The model differentiates between unique genome-wide (black) and nonsynonymous (orange) rates for each individual, while applying a single synonymous rate (green) across all individuals. **(b)** Illustrates the evolutionary distance over time for three selected persistently infected individuals – see Supplementary Figure 7 for all 576 persistent infections. Points on the graphs represent the total genetic distance from the consensus sequence at the initial time point, calculated based on allele frequency changes over time. Dashed lines indicate the regression lines that best fit these data. **(c)** Shows the mutant allele frequency trajectories for the three persistent infections examined, categorised into synonymous, nonsynonymous, and non-coding (grey) mutations – see Supplementary Figure 2 for trajectories in all individuals with measurable evolution in at least 3 time points. Each mutation that reached a minimum frequency of 20% at least at one time point is shown. We can see partial and full sweeps of de novo mutations over the course of persistent infections. A horizontal grey line across the graphs marks the 20% allele frequency threshold.

The considerably higher rate of nonsynonymous evolution indicates at least some nonsynonymous mutations are subject to positive selection, and moreover that this selective pressure differs among individuals. In contrast, the preference for a regression model with a single rate for synonymous mutations implies that these mutations are evolutionarily neutral or nearly neutral, evolving at approximately the same rate across all individuals.

To assess how well our model choices fit the data, we further examined the 13 persistent infections with three or more sequenced samples, and that included at least one measurement of genetic distance for both synonymous and nonsynonymous mutations (Supplementary Figure 2). The best fit regression lines captured most of the changes in the genetic distances over time, with nonsynonymous mutations occurring more frequently, and reaching higher frequencies, than synonymous ones (Supplementary Figures 2a,b; see also Figures 5b**,c**). We typically observed two distinct patterns in allele frequencies across different persistent infections. In some cases, transient alleles emerged together at one time point, before disappearing at the later time points, suggesting we are capturing distinct subpopulations within infections. In other cases, we observed the near complete sweep of mutations from low to high frequencies. Other cases were largely a combination of both patterns with some mutations appearing and disappearing in groups while others were present throughout most of the infection (see Supplementary Figure 3).

### The nonsynonymous divergence rate is highest in the receptor binding domain

To explore evolutionary rate variation across the genome, we next assumed the consensus sequence at baseline represents the founder virus, and that the start of infection occurred at the midpoint between the last negative PCR test and the first sampled time point of the persistent infection. For the majority of infections, the last negative PCR test was taken between 20 to 40 days before the baseline (see Supplementary Figure 4). Using the estimated infection start dates, we calculated an evolutionary rate for each region of the genome, aggregating across all individuals (see **Methods**). We called this the divergence rate to distinguish it from the approach we took to measure evolutionary rates per individual, because most infections had only a limited number of mutations, which precluded a calculation of a per-individual rate per gene or gene segment. This commonly used approach to measuring within-host divergence rates comes with two key disadvantages compared to the intra-infection evolutionary rates we measured in the previous section. First, it requires estimating the time elapsed since the start of the infection rather than using only known sample collection dates. Second, this method has a tendency to ascribe any changes in allele frequencies, or their absence, to substitution rates rather than to sampling noise.

We observed considerable variability in the rate of divergence across the genome (Figure 6). The bulk of this rate variation among different genomic regions came from nonsynonymous changes, with the rate of synonymous divergence remaining relatively uniform across most regions, except for the *M* and *N* genes which had a synonymous rate nearly double that of the other regions (Figure 6a). *ORF8* and *S* had the highest rates of nonsynonymous divergence, nearly five times greater than the rates of synonymous divergence, whereas *ORF6* showed the lowest rate of nonsynonymous divergence, further indicating it is likely under strong purifying selection.

**Figure 6:**
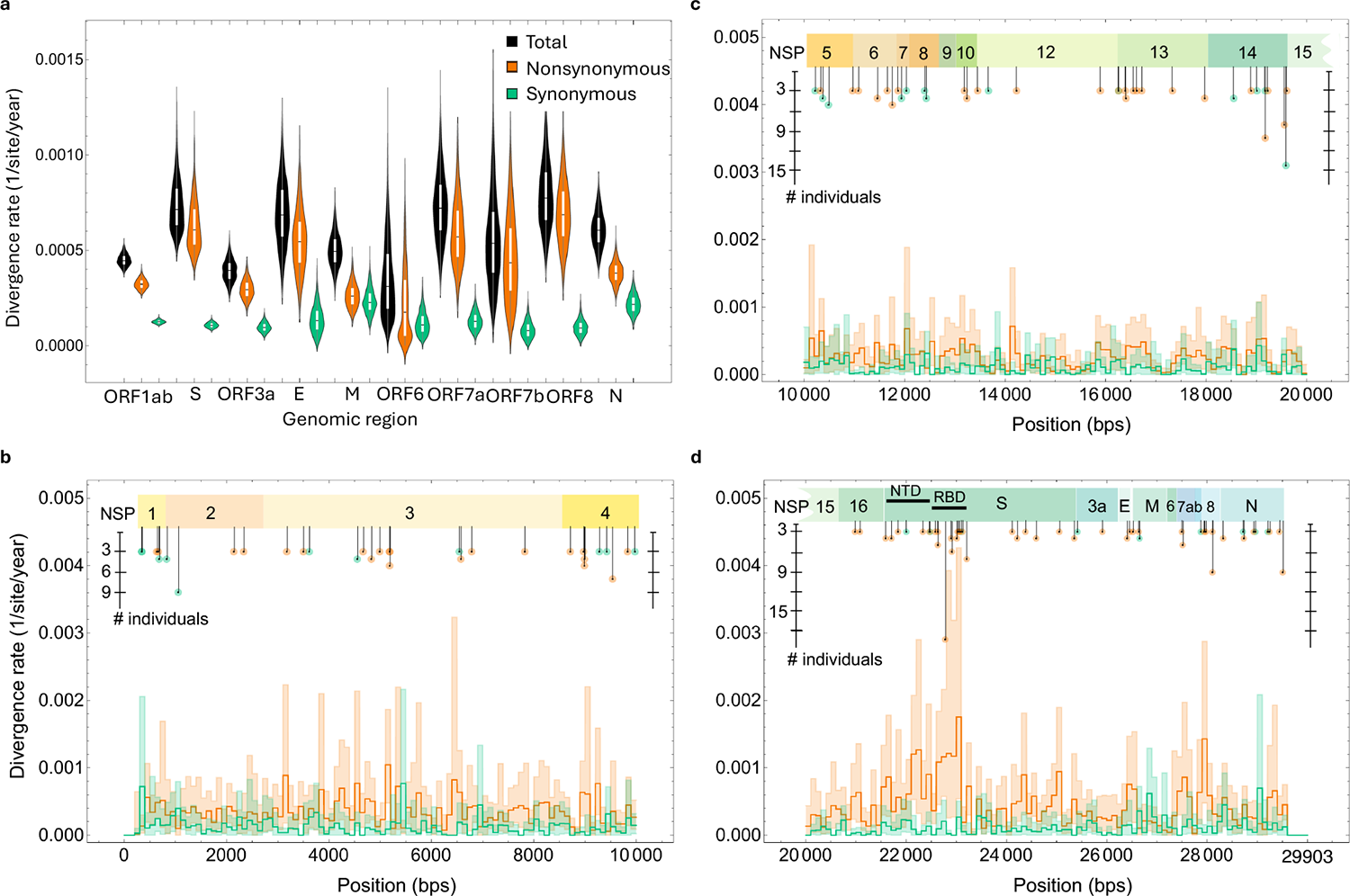
Virus divergence rates across the genome. **(a)** This panel presents the estimated divergence rates from the putative founder, showcasing genome-wide (black), nonsynonymous (orange), and synonymous (green) substitution rates across different regions. The distributions represent the bootstrap estimates derived from 576 persistent infections. **(b-d)** Display the estimated divergence rate per 100 (nonoverlapping) base pair segments of the genome for non-structural proteins (NSPs): NSP 1 to 4 in (b), NSP 5 to 15 in (c), and NSP 15 and 16, along with other structural non-structural proteins and accessory factors in (d). Shaded area represents the 95% confidence intervals from bootstrapping. Recurrent mutations identified in three or more persistent infections are highlighted.

Looking at divergence rates across non-overlapping gene segments of 100 base pairs in length, most segments in *ORF1ab* and *S*, which together make up approximately 85% of the SARS-CoV-2 genome, displayed low levels of variation in synonymous divergence rates, while nonsynonymous rates varied up to 5 times in some segments of *ORF1ab*, and 10 times in *S* (Figure 6b-d). The end tail of the RBD in *S* (22990 to 23090) had the highest rates of nonsynonymous divergence, suggesting that it is under strongest positive selection (Figure 6d). Accelerated nonsynonymous evolution in the NTD and RBD during persistent infections supports the idea that these infections are the main source behind the emergence of highly divergent variants at the population level. New major lineages that successfully spread also exhibit an overabundance of nonsynonymous mutations in the same genomic regions compared to other circulating lineages at the time of their emergence in the population.

### Recurrent within-host mutations with transient fitness advantage

We found 379 (262 nonsynonymous and 117 synonymous) mutations found in at least two individuals among the 576 persistent infections (**Source file**; see also Figure 5b-d). The highest concentration of these recurrent mutations that were nonsynonymous were in *ORF8* (24 mutations), *E* (14 mutations), and *S* (210 mutations), whereas the highest concentration of recurrent synonymous mutations was in *ORF7b* (3 mutations) and *M* (14 mutations).

The per-lineage fitness effect of recurrent mutations was measured at the between-host level using a globally representative SARS-CoV-2 phylogeny ^25^. When fitness effects were examined within the same major lineage as the virus from persistent infections, 54% of these mutations showed a positive fitness effect (Supplementary Figure 5a-c). Most recurrent mutations also had very low population-prevalence with nearly 47% being present in less than 0.01% of all samples within ONS-CIS sequences from the same major lineage as the virus from persistent infections (Supplementary Figure 5d). This suggests that almost half of the recurrent mutations have a fitness advantage at the within-host level but a fitness disadvantage and low prevalence at the between-host level.

The most recurrent mutations were S:N405D (with corresponding nucleotide substitution A22775G in 8 infections), NSP14: T516T (T19587A, in 13 infections), and NSP14:C382G (T19183G, in 10 infections), all of which were found in persistent Omicron infections, BA.2, BA.4, and BA.5. The highly recurrent *Spike* mutations that were found in at least three persistent infections and had very high between-host fitness effects were S:L452R, S:K356T, and S:T547K all of which are lineage-defining mutations (see Supplementary Figure 5e). In particular, S:K356T is lineage-defining for BA.2.86 and was found in multiple BA.2 and BA.5 persistent infections. On the other hand, most of the highly recurrent mutations with strong negative between-host fitness effects were concentrated in various non-structural proteins of *ORF1ab* (see Supplementary Figure 5e).

We also investigated potential associations between host characteristics and recurrent mutations in SARS-CoV-2 persistent infections. Specifically, we examined whether there is an association between the age group of the persistently infected individual and the number of times a mutation recurs (Supplementary Figure 5f), the between-host fitness effect of recurrent mutations and the age group of the individual in which they appeared (Supplementary Figure 5g), and the fitness effect of the recurrent mutations with respect to the duration of persistent infections (Supplementary Figure 5h). However, we found no strong associations between these factors.

### Infection duration is correlated with evolutionary rates

We found no significant associations (ΔBIC < 0) of age, sex, vaccination status, prior infection, or virus lineage with within-host evolution rates. This evaluation was based on comparing the BIC values of the best-fit regression model for determining within-host rates with models that included each of these parameters as an additional fixed effect (see **Supplementary Table 2**). Notably, our observation that the within-host evolutionary rates do not significantly differ between vaccinated and unvaccinated individuals suggests that vaccination does not lead to accelerated evolutionary rates.

We were also interested in investigating whether experiencing a viral rebound had an impact on evolutionary rates. To do this, we categorised persistent infections into either persistent-chronic (consistently positive PCR tests throughout the infection) or persistent-rebounding (at least one negative PCR test during the infection); see also ref ^8^ for more about these two categories. We found weak evidence (ΔBIC=1) in support of a positive association between experiencing a rebounding viral load and an elevated nonsynonymous evolutionary rate. After controlling for duration of infection, since it is more likely to identify persistent-rebounding infections when the infections are longer (i.e. more time to pick up a negative PCR test during a prolonged infection), by only examining a subset of infection where the duration of infection is longer than at least 56 days, we found no association between viral rebound and higher evolutionary rates (ΔBIC <0). However, we did identify a positive association (ΔBIC >2) between the evolutionary rates and the duration of infection, indicating that longer infections exhibit higher rates of nonsynonymous evolution. To determine if this association was biased by the lower genetic diversity typically seen in shorter infections, which could result in lower evolutionary rate estimates, we also examined longer infections lasting at least 56 days. Our analysis confirmed statistical support (ΔBIC >2) for the positive relationship between infection duration and evolutionary rates, even within these subsets of infections (see **Supplementary Table 2**).

## Discussion

We characterised viral genomic diversity and within-host evolutionary rates in 576 individuals with persistent SARS-CoV-2 infections, identified through large-scale community surveillance, and including samples collected between November 2020 to March 2023. Central to our investigation was the hypothesis that persistent infections could serve as the primary source for the saltatory evolution of the virus at the between-host level, mirroring the same evolutionary changes we see with the emergence of highly-divergent variants. This premise led us to identify host characteristics associated with prolonged infections and to characterise viral evolutionary patterns across the genome and between individuals.

We observed significant variability in within-host viral evolutionary rates between infections. This variability was predominantly attributed to the different rates at which individuals accumulated nonsynonymous mutations, with the rate of synonymous mutations being similar among all individuals and typically more than four-fold slower than the rate of nonsynonymous mutations. This variability among individuals explains previous findings of limited consensus change mutations in some individuals and over-abundance of mutations in others ^8,26^. We also observed considerable variability in nonsynonymous evolutionary rates across most of the genome, but not synonymous rates, with the receptor binding domain of the Spike protein having the highest rate of nonsynonymous evolution relative to all other genomic regions. We also found elevated synonymous rates in *M* and *N* genes which suggest they could have functional benefit for mRNA stability and translation efficacy, particularly on phosphorylation sites that are abundant in *N* ^9^.

Although older individuals were more likely to experience persistent infections, we found no evidence to suggest that host factors such as age, sex, vaccination status, virus lineage, previous infection, or dynamics of viral RNA titres significantly affected evolutionary rates. However, we did observe a positive association between evolutionary rates and the duration of infection, with longer-lasting infections exhibiting higher rates of nonsynonymous evolution. We speculate that individuals with longer infections may have more impaired immune responses, and/or be undergoing treatment, which may result in faster rates of adaptive evolution. Our examination of recurrent within-host mutations which are rare in the general population and have negative between-host fitness effects further illustrates the complex evolutionary dynamics at play within persistent infections. These mutations likely confer a selective advantage within hosts due to enhanced replication rates and/or immune evasion. However, they may prove detrimental at the between-host level, for example if they result in reduced transmissibility of the virus between individuals ^11,27,28^.

We found that *ORF6* had the lowest levels of nonsynonymous diversity and divergence rate compared to the other genomic regions, indicating it is functionally conserved during persistent infections. Strikingly, we found no diversity in the second codon position of *ORF6*; all mutations at this position would be nonsynonymous. These observations are consistent with several studies that have highlighted the crucial role of *ORF6* in viral replication and disease progression ^29–31^. These results suggest that *ORF6* could be a promising candidate for the development of therapeutic drugs for treating individuals with persistent infections^32^.

Many of the recurrent mutations identified in our study have been found to have functional importance for SARS-CoV-2. For example, the mutation S:G446V is linked to treatment resistance ^33^. The mutation NSP3:T820I frequently occurs in patients treated with Nirmatrelvir and Ritonavir ^34^, while NSP7:L3935L is commonly found in cancer patients and those undergoing immunosuppressive or steroid therapies ^35^. Another mutation, S:D1153Y, is known for its antibody escape properties ^36^. The mutation M:N117K may play a role in the glycosylation of the virus ^37^. Also, recurrent mutations S:L216F, S:S98F, and N:P151S have previously been identified as being under multilevel selection, beneficial at the within-host level but deleterious at the between-host level ^11^.

Our findings shed light on the complex interplay between persistent SARS-CoV-2 infections, the demographic characteristics of those infected, and the evolutionary mechanisms driving the virus evolution within these individuals. This study also underscores how persistent infections may contribute to the emergence of highly divergent variants, with factors such as the duration of infection and accelerated rate of evolution at nonsynonymous sites, particularly in the RBD of Spike protein, influencing their evolutionary rates.

## Methods

### ONS COVID-19 Infection Survey

This work contains statistical data from ONS which is Crown Copyright. The use of the ONS statistical data in this work does not imply the endorsement of the ONS in relation to the interpretation or analysis of the statistical data. This work uses research datasets which may not exactly reproduce National Statistics aggregates.

The Office for National Statistics Covid-19 Infection Survey (ONS-CIS) is a UK household-based surveillance study, which began in the UK from April 2020 ^38^ and was first paused in March 2023 ^2^. Our analysis here covered the period from 2 Nov 2020 to 21 March 2023. Households from nationwide address lists were invited to participate (every household member aged two years and above), ensuring as representative a cross-section of the population as possible. Participants gave written informed consent to contribute swab samples (self-collected or by a parent/carer for those under 12 years), irrespective of symptoms, and completed a questionnaire for each assessment.

Most of the participants in the survey consented to routine PCR sampling at weekly intervals for the first month of enrollment and monthly thereafter for the duration of the study ^2,8^. From December 2020, all cases where a participant tested positive with a high viral load (Ct ≤30), their sample was further sent for sequencing.

### Sequencing

Samples were sequenced at one of five sequencing centres, University of Oxford (OXON), Northumbria University and associated NHS foundation trusts (NORT), National Infection Service Public Health England (PHEC), Quadram Institute Bioscience, Norwich (NORW), and Wellcome Sanger Institute (Sanger). The great majority of samples were sequenced on Illumina Novaseq, with the rest using Oxford Nanopore GridION or MINION. The standard consensus FASTA sequences for all ONS-CIS samples were generated using the ARTIC Nextflow processing pipeline (v1) ^39^, or veSeq, an RNA sequencing protocol based on a quantitative targeted enrichment strategy ^2,40^ with consensus sequences produced using Shiver (v1.5.8) ^41^. For additional information about the survey, sequencing protocol, and FASTA consensus sequence protocol see ^2,8^.

### Identification of persistent infections

We used the consensus sequences generated using ARTIC Nextflow or Shiver to determine whether two or more sequences from the same individual were from the same infection, using the method outlined in ^8^. Briefly, if two sequences from the same individual were collected at least 26 days apart, were of the same major lineage, and shared a rare single nucleotide polymorphism (SNP) compared to the population-level consensus, the individual was determined to be persistently infected. Our analysis covered infections with the Alpha, Delta, Omicron BA.1, BA.2, BA.4, BA.5, and XBB major lineages, and a SNP was deemed to be rare if found in <400 samples of that lineage (see Supplementary Figure 6). Due to possible misclassification of some BA.2 sequences as BA.5 and vice versa using the Pango lineage nomenclature ^42^, we considered the possibility that some BA.5 sequences could belong to a BA.2 infection. This approach identified 3 cases of BA.2 persistent infections, which included at least one sequence misclassified as a BA.5 lineage. Without requiring any additional adjustment to separate second-generation BA.2 (e.g. BA.2.75) and BA.5 (e.g. BQ.1) major lineages from their closest ancestors, our method reliably recovered subsets of infections within BA.2 and BA.5 that were attributable to second-generation variants. Specifically, we found 21 BA.2.75 and 25 BQ.1 persistent infections.

### Identifying intra-host single nucleotide variants

We called an intra-host single nucleotide variant (iSNV) at a given position in the genome if there were 10 or more bases called at that position, including gaps, and if the most common minor allele was present at 20% or more but less than 50%. The small number of bases required to call an iSNV was chosen because many samples had low viral titre, whilst the 20% threshold was to avoid biases introduced by differing amounts of sequencing noise across all the samples.

We also identified mutations, which we defined as iSNVs or major alleles that differed from the majority allele at the first sampling time point, and reached at least 20% frequency at baseline or any of the subsequent time points. Whereas iSNVs are always less than 50% frequency by definition, a mutation can be at any frequency above 20% (including 100%). To ensure consistency of methods across our analyses, we also defined the majority-rule consensus at each sampling as the majority allele, with a minimum of 10 bases to call a consensus at any given position. Unless stated otherwise, when we refer to the consensus we mean the majority-rule consensus, not the consensus generated using ARTIC Nextflow or Shiver.

Some positions in the genome are prone to having low frequency iSNVs in a high proportion of samples, and are often sequencing centre specific. Although we do not know what causes these low frequency iSNVs, they are unlikely maintained through descent and we therefore label them ‘artefactual iSNVs’. For each sequencing centre in our study, we masked genomic positions where an iSNV was present at ≥2% frequency in more than 1% of samples from that sequencing centre.

### Nucleotide diversity

Nucleotide diversity was calculated using the *π* statistic, which is the common measure of diversity least affected by the number of sequences used in the analysis ^43^. For each persistent infection, nucleotide diversity at a given time point is given by:

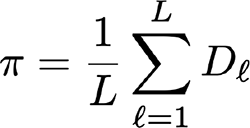

where *L* represents the number of nucleotide positions being examined, and *D_l_* the genetic diversity at locus *l* with an iSNV present at a frequency ≥20%. This is calculated as:

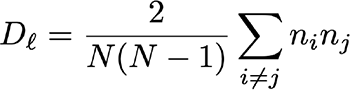

where *n_i_* represents the number of nucleotides *i* = A, C, G or T (not including gaps), and *N* the total number of reads at that locus.

### Estimating within-host genetic distance

We used differences in mutant allele frequencies between two sequences from the same infection to calculate the genetic distance between the sequences. This is similar to an approach that has been used to measure within-host evolutionary rates of influenza A in a chronically infected individual ^24^. We calculated changes in allele frequency relative to the first sequenced time point in each persistent infection.

Synonymous and nonsynonymous distance was determined by whether the mutant allele would result in the same (synonymous) or a different (nonsynonymous) amino acid being coded for compared to the first time point in the infection.

Following this definition of evolutionary distance, a mutant allele *i*, present at frequency *f_i_*(t_0_) at the first time point and *f_i_*(t*_k_*) at the *k*th time point contributes |*f_i_*(*t_k_*) - *f_i_*(*t*_0_)| to the pairwise distance between the two sequences. More generally, if the pair of sequences has *M* mutant alleles, the total genetic distance between them is

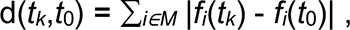

where |.| represents the absolute change of allele frequency. We excluded pairs of samples where the total number of overlapping base pairs between the two consensus sequences is smaller than 50% of genome length as these can give rise to deflated or inflated measures of genetic distance per site.

### Estimating within-host evolutionary rate

We quantified within-host evolutionary rates by assuming a linear relationship between the genetic distance and the time elapsed since the first sequence was collected from each individual.

A linear regression model represented the changes in genetic distance relative to first sequence over time within each persistent infection as

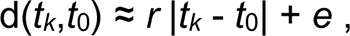

where *r* is the evolutionary rate and *e* is the y-intercept, which represents the expected amount of noise when measuring genetic distance. The noise could arise from either sequencing error or undiagnosed population structure ^24^. If a persistent infection does not have a detectable mutant allele that reaches frequency ≥20%, we exclude that individual from evolutionary rate analysis as we cannot quantify the contribution of noise in frequency change of alleles.

Our analysis encompassed five different regression models with varied levels of complexity (see **Supplementary Table 1**) to estimate genome-wide, synonymous, and nonsynonymous within-host evolutionary rates. We used the Bayesian Information Criterion (BIC) value for model selection, balancing model complexity against fit quality.

The y-intercept can be interpreted as the baseline level of noise in changes of allele frequencies. With a fixed nonsynonymous y-intercept at 3.4×10^-5^ substitutions per site and an average of 4.5 nonsynonymous mutations per infection, we can estimate that roughly 23% of the variations in nonsynonymous allele frequencies may be attributed to noise. Conversely, for a typical synonymous mutation characterised by a y-intercept of 2.2×10^-5^ substitutions per site and an average of 1.6 synonymous mutations per infection, about 40% of changes in allele frequencies are driven by noise. While we expect the contribution of sampling noise to be the same for both synonymous and nonsynonymous mutations, biological factors such as selection and functional constraints may not be uniform across different mutation types. More specifically, given that synonymous mutations are more likely to be neutral or nearly neutral, their baseline noise can be more reflective of sampling noise and the stochastic nature of viral replication and mutation.

We examined the following linear regression models for measuring evolutionary rates and baseline noise:

i. Complete pooling: *d_i_*(*t*) = *r_0_ t* + *e_0_* + *ε_i_*(*t*) This model assumes a single (fixed) underlying rate, denoted as *r_0_*, and intercept, *e_0_*, which describes a common evolutionary rate and noise contribution across all individuals. The error term *ε_i_*(*t*) represents the residual unexplained variability in distance, *d_i_*(*t*) for persistent infection *i*. Models (ii) to (v) all incorporate partial pooling with varying degrees of complexity.
ii. (ii) Random intercept: *d_i_*(*t*) = *r_0_ t* + *e_i_* + *e_0_* + *ε_i_*(*t*) A linear mixed effect model which assumes a shared rate, *r_0_*, and error, *e_0_*, across all infections (fixed effects)with each infection *i* also having a unique intercept *e_i_*, indicative of individual-level noise variation (random effect).
iii. (iii) Random slope with one fixed intercept: *d_i_*(*t*) = (*r_0_+r_i_*) *t* + *e_0_* + *ε_i_*(*t*) A linear mixed effect model which assumes a single (fixed) underlying rate,*r_0_*, and error, *e_0_*, shared by all individuals in addition to a unique underlying rate, *r_i_*, for each persistent infection, *i* (random effect).
iv. (iv) Random slope with multiple fixed intercepts: *d_i_*(*t*) = (*r_0_+r_i_*) *t* + *∑_j_ e_j_* + *ε_i_*(*t*) Considering potential sequencing centre-specific noise, we categorised y-intercepts into nine groups, based on where the sequences were sampled. For instance, if the initial sample from a persistently infected individual was sequenced in Sanger Institute (“Sanger”) and a subsequent sample in the University of Oxford (“OXON”), the y-intercept corresponding to this persistent infection belong to the *j*=(“Sanger”, “OXON”) category. There are a total of nine such y-intercept categories, represented as j∊{(NORT, PHEC), (NORT, NORW), (NORT, Sanger), (OXON, PHEC), (Sanger, OXON), (NORT), (PHEC), (OXON), (Sanger)}. There are 9 pairs of samples that are (NORT, PHEC), 4 (NORT, NORW), 90 (NORT, Sanger), 14 (OXON, PHEC), 1 (Sanger, OXON), 147 (NORT), 16 (PHEC), 10 (OXON), and 331 (Sanger). We assessed these categories for their impact on baseline noise in the data, assuming their influence is constant over time. This model therefore introduces nine fixed effects *e_j_* to account for variations in y-intercepts due to sequencing noise levels.
v. _(v)_ No pooling: d*_i_*(*t*) = *r_i_ t* + *e_i_*

Each persistent infection, denoted as *i*, has a unique rate and error term. In practice, this model cannot be applied to our dataset because the number of measurements is smaller than the number of random effects, as persistent infections with only two samples yield a single measurement for genetic distance.

Our analysis showed, based on the lowest BIC value, that the random slope with one fixed intercept regression model (iii) best explains genome-wide and nonsynonymous evolutionary rates while the random intercept regression model (ii) best explains synonymous rate for persistent infections. The lines of best fit for all the persistent infections with measurable evolution is shown in Supplementary Figure 7.

### Estimating within- and between-lineage rates at the between-host level

To assess the saltatory evolution of SARS-CoV-2 at the between-host level, we used a previously identified representative sample from the ONS-CIS dataset ^2^. This dataset covered sequences from the Alpha, Delta, Omicron BA.1, BA.2 (excluding BA.2.75), BA.2.75, BA.4, BA.5 (excluding BQ.1), and BQ.1 lineages. We then constructed the ancestral sequence for each major lineage using TreeTime ^44^ and calculated total, nonsynonymous, and synonymous Hamming distances between samples from each major lineage relative to the ancestral sequence of the same major lineage. Finally, to estimate the between-lineage rate, we calculated the total, nonsynonymous, and synonymous Hamming distances between the Wuhan reference sequence (NC_045512.2) and the ancestral sequence for each major lineage.

### Divergence rate from putative founder

Since persistent infections on average have 5 mutations across the genome (IQR: 2, 8), estimating an evolutionary rate for different segments of the genome at an individual level is not practical. We therefore used the majority-rule consensus sequence at the first time point of each persistent infection as a proxy for the founding virus. We then estimated the start time of infection as the midpoint between the last negative PCR test and the first sequence from the persistent infection. We measured the typical evolutionary rate (rather than mean) from the putative founder across all individuals for each segment of the genome.

While this method is frequently used for calculating within-host divergence rates for viruses like HIV ^45^, it will miss early fixation events that might have shifted the consensus sequence away from the true founding virus by the time the first sample was collected; assumes the founding viral population was genetically homogeneous ^46^; does not control for noise which could bias estimates of the divergence rate. Nonetheless, aggregating across a large number of individuals should help mitigate these effects.

This approach involved treating each measurement of divergence from the putative founder at any given time point, *t*, as an independent observation, regardless of its associated persistent infection. The divergence from the founder for each genomic segment at any time point, including baseline, was defined as the cumulative frequency of all mutant alleles within that segment at time *t*. For example, if there were no mutant alleles within a genomic segment at a given time point, we recorded a divergence of zero. Subsequently, we used a linear regression with a zero y-intercept at the start time of infection to calculate the divergence rate from the putative founder for each genomic segment. This can be expressed as *d^(n)^_i_*(*t*) = *r_i_ t* + *ε^(n)^_i_*(*t*), where *r_i_* is the divergence rate for genomic segment *i*, and *d^(n)^_i_*(*t*) is calculated as the genetic divergence of sample *n* from its putative founder within segment *i* at time *t*. Each sample, *n*, from a persistent infection represents one measurement of *d^(n)^_i_*(*t*). If a sample is collected at time *t=t\** and has no mutant alleles within segment *i*, then *d^(n)^_i_*(*t\**)=0. For each sample, the estimated start of infection is taken as *t=*0. Each sample from an individual acts as an independent observation of genetic distance for segment *i*. The error term *ε^(n)^_i_*(*t*) represents the residual unexplained variability in distance, *d^(n)^_i_*(*t*), for sample *n*.

To ensure an equal representation of each persistent infection in the divergence rate assessment for a genomic segment, we limited our analysis to two divergence measurements per individual—one at the baseline and another selected randomly from later in the infection. We then performed bootstrapping across all individuals and every possible pair of divergence measurements per individual to create a distribution of divergence rate estimates for each genomic segment.

## Data availability

All raw consensus sequences have been made publicly available as part of the COG-UK Consortium (https://webarchive.nationalarchives.gov.uk/ukgwa/20230505214946/https://www.cogconsortium.uk/priority-areas/data-linkage-analysis/) and are available from the European Nucleotide Archive at EMBL-EBI under accession number PRJEB37886.

## Acknowledgement

The CIS was funded by the Department of Health and Social Care and the UK Health Security Agency, with in-kind support from the Welsh Government, the Department of Health on behalf of the Northern Ireland Government and the Scottish Government. The COVID-19 Infection Survey Group of the COVID-19 Genomics UK (COG-UK) Consortium was supported by funding from the Medical Research Council part of UK Research & Innovation, the National Institute of Health Research (NIHR) (grant code: MC_PC_19027) and Genome Research Limited, operating as the Wellcome Sanger Institute. We acknowledge use of data generated through the COVID-19 Genomics Programme funded by the Department of Health and Social Care. A.S.W. is supported by the NIHR Health Protection Research Unit in Healthcare Associated Infections and Antimicrobial Resistance at the University of Oxford in partnership with the UK Health Security Agency (NIHR200915) and the NIHR Oxford Biomedical Research Centre, and is an NIHR Senior Investigator. K.L. is supported by the Royal Society and the Wellcome Trust (107652/Z/15/Z) and by the Li Ka Shing Foundation. The research was supported by the Wellcome Trust Core Award grant number 203141/Z/16/Z, with funding from the NIHR Oxford BRC. The views expressed are those of the authors and not necessarily those of the NHS, the NIHR, the Department of Health, the Department of Health and Social Care or the UK Health Security Agency.

## Supplementary Tables

**Supplementary Table 1:**
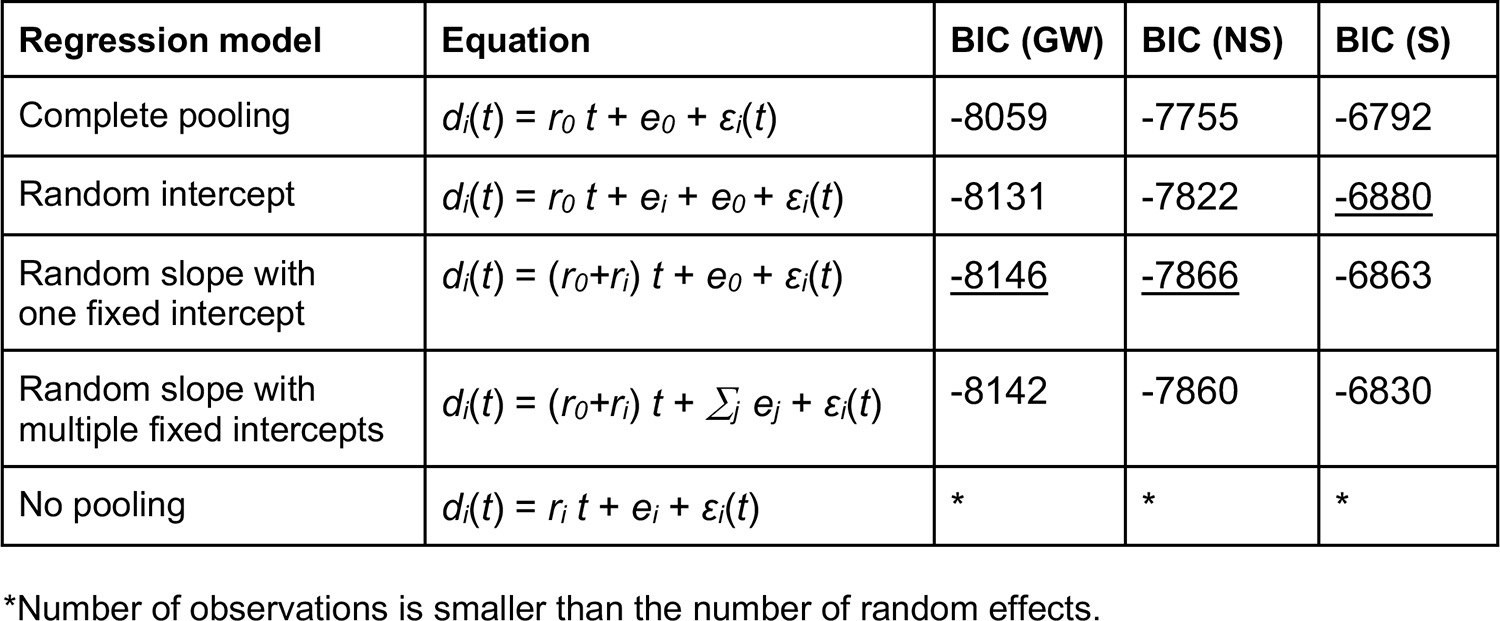
Model comparison for estimating within-host evolutionary rates. Comparison of regression models for estimating genome-wide (GW), nonsynonymous (NS), and synonymous (S) evolutionary rates. Each model is presented with its corresponding equation and Bayesian Information Criterion (BIC) value, which assesses model fit to the data. Parameters *e_0_* and *r_0_* represent fixed effects for y-intercept at time *t*=0 (corresponding to the day when the first sample from a persistent infection was collected) and rate across all persistent infections, respectively; *d_i_*(*t*) represents distance at time *t* for persistent infection *i* (dependent variable); *r_i_* and *e_i_* represent random effects for evolutionary rate and intercept per persistent infection, respectively; *ε_i_*(*t*) is the error term which represents the unexplained variability in the dependent variable; the index *j* corresponds to nine categories for y-intercept labelled based on sequencing centre(s) that genetic samples are collected from. Models with lowest BIC values are highlighted with an <u>underline</u>.

| Regression model | Equation | BIC (GW) | BIC (NS) | BIC (S) |
| --- | --- | --- | --- | --- |
| Complete pooling | $d_i(t) = r_0 t + e_0 + \varepsilon_i(t)$ | -8059 | -7755 | -6792 |
| Random intercept | $d_i(t) = r_0 t + e_i + e_0 + \varepsilon_i(t)$ | -8131 | -7822 | <u>-6880</u> |
| Random slope with one fixed intercept | $d_i(t) = (r_0 + r_i) t + e_0 + \varepsilon_i(t)$ | <u>-8146</u> | <u>-7866</u> | -6863 |
| Random slope with multiple fixed intercepts | $d_i(t) = (r_0 + r_i) t + \sum_j e_j + \varepsilon_i(t)$ | -8142 | -7860 | -6830 |
| No pooling | $d_i(t) = r_i t + e_i + \varepsilon_i(t)$ | * | * | * |

\*Number of observations is smaller than the number of random effects.

**Supplementary Table 2:** Evaluation of associations between various host factors and within-host evolutionary rates. **(a)** This table examines the impact of integrating individual host factors—age, sex, vaccination status, prior infection, virus lineage, duration of infection, and RNA viral load dynamics—into the best-fit regression model as fixed effect parameters and comparing best fits using the Bayesian Information Criterion (BIC) values. The baseline model is a linear mixed-effects regression, identified as the optimal fit for genome-wide (GW), nonsynonymous (NS), and synonymous (S) distances over time (see Supplementary Table 1). Each of the seven factors is added as a fixed effect to this baseline model, with categorical variables including age (aged 60 and above: 295; aged below 60: 199), sex (male: 293; female: 201), vaccination status (received at least one dose: 470; no vaccination: 24), prior infection (none: 478; at least one: 16), viral lineage (10 Alpha, 95 Delta, 87 BA.1, 173 BA.2, 14 BA.4, 111 BA.5, and 4 XBB with measurable evolution), and viral load dynamics (experienced viral rebound: 32; no rebound detected: 462). Duration of infection is classed as a continuous variable ranging from 26 to 316 days per infection. **(b)** Comparing the BIC values for a subset of infections with durations lasting longer than 36 days (198 infections) and 56 days (110 infections) between the null model and a model that includes duration of infection as an additional fixed effect parameter.

**(a)**
| Fixed effects | BIC (GW) | BIC (NS) | BIC (S) |
| --- | --- | --- | --- |
| Null model | -8146 | -7866 | -6880 |
| Virus lineage | -8120 | -7838 | -6847 |
| Prior infection | -8141 | -7860 | -6874 |
| Vaccination status | -8141 | -7861 | -6874 |
| Sex | -8142 | -7862 | -6874 |
| Age | -8143 | -7861 | -6874 |
| Viral load dynamics | -8145 | -7867 | -6877 |
| Duration of infection | -8150* | -7868* | -6881 |

**(b)**
| Fixed effects | BIC (GW) | BIC (NS) | BIC (S) |
| --- | --- | --- | --- |
| Null model (t>36) | -2907 | -2925 | -2661 |
| Duration of infection (t>36) | -2910* | -2928* | -2659 |
| Null model (t>56) | -1559 | -1558 | -1605 |
| Duration of infection (t>56) | -1560 | -1560* | -1602 |

\*Indicates $\Delta\text{BIC} = \text{BIC}_{\text{Null}} - \text{BIC}_{\text{Alternative}} > 2$ .

### Supplementary Figures

**Supplementary Figure 1:**
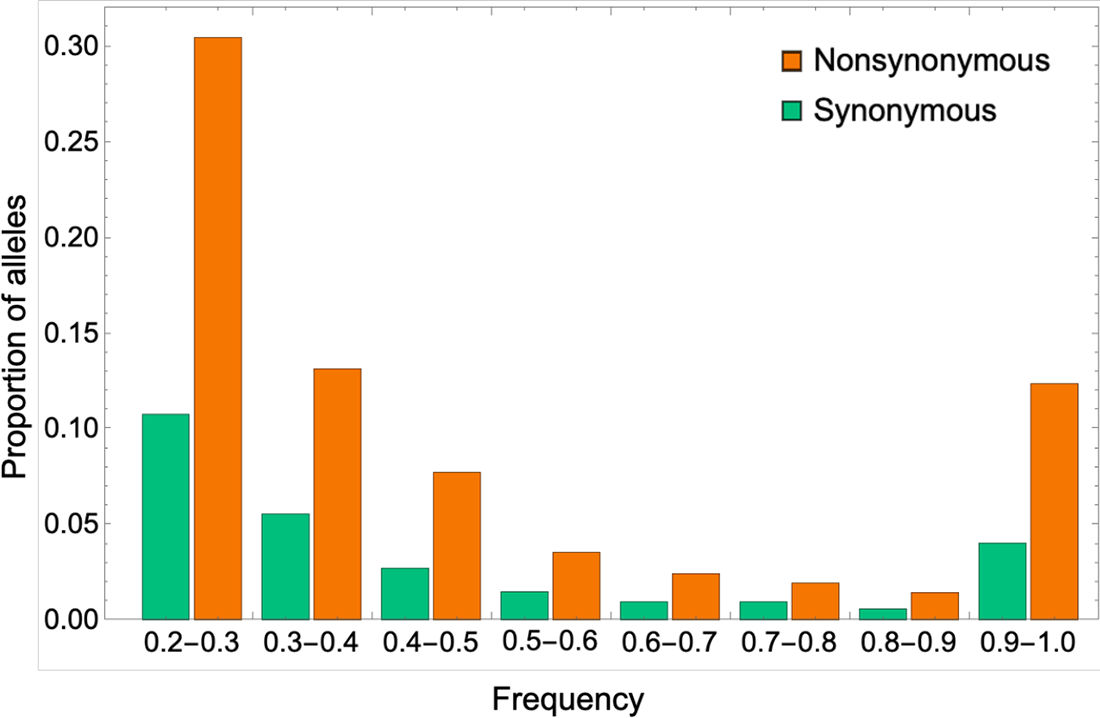
Site frequency spectrum. Proportion of synonymous (green) and nonsynonymous (orange) mutations in persistent infections across all frequency bands.

**Supplementary Figure 2:**
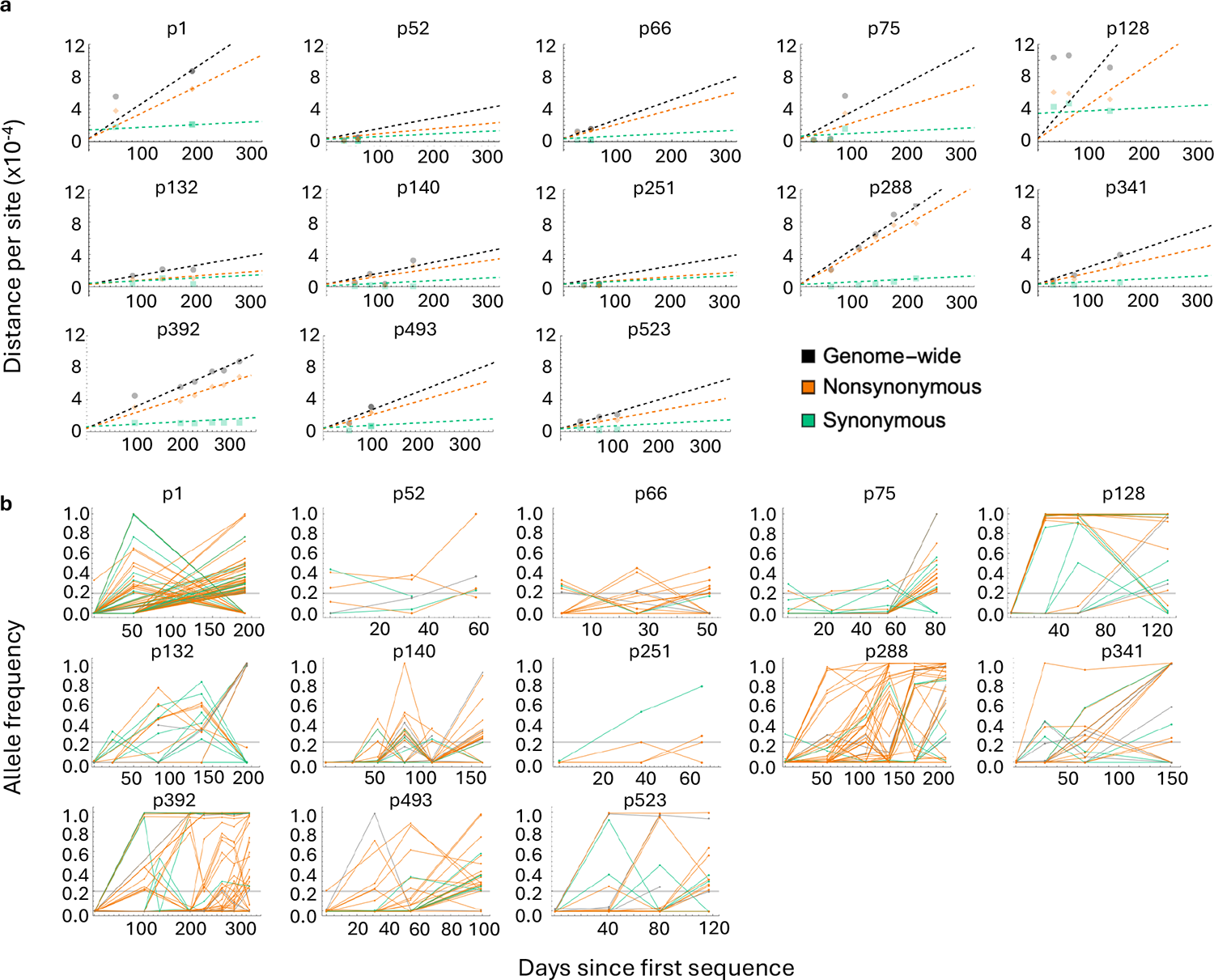
Rates of genome-wide, nonsynonymous, and synonymous evolution in 13 persistently infected individuals. **(a)** Illustrates the evolutionary distance over time for a subset of 13 persistently infected individuals, each characterised by a minimum of three temporal data points and the presence of at least one synonymous and one nonsynonymous mutant allele. Points on the graph represent the total genetic distance from the consensus sequence at the initial time point, calculated based on allele frequency changes over time. Dashed lines indicate the regression lines that best fit these data. **(c)** Shows the allele frequency trajectories for the 13 persistent infections examined, categorised into synonymous, nonsynonymous, and non-coding (grey) mutations. Each mutation that reached a minimum frequency of 20% at least at one time point is shown. A horizontal grey line across the graphs marks the 20% allele frequency threshold.

**Supplementary Figure 3:**
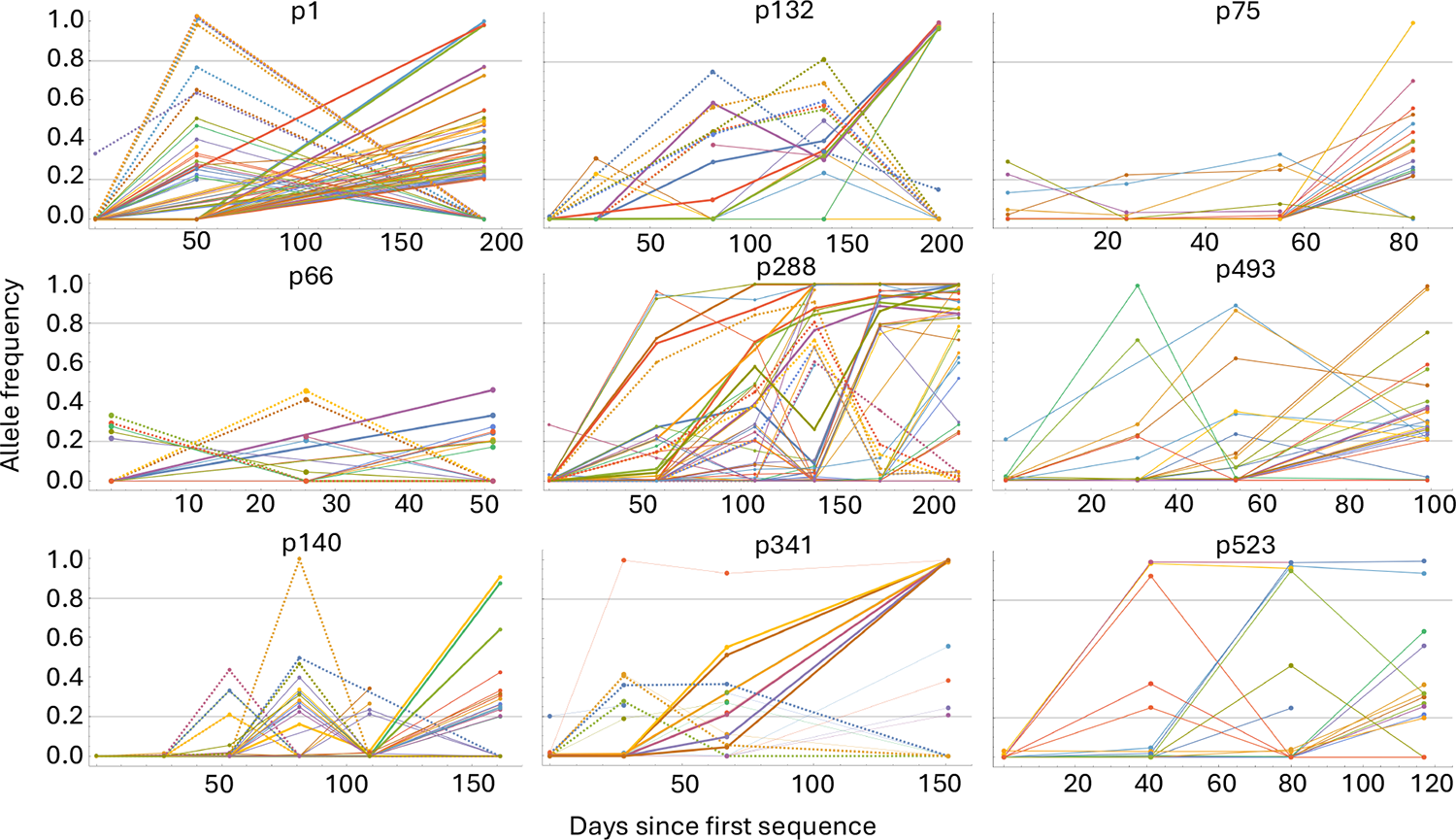
Temporal allele frequency dynamics in nine persistent infections. The figure illustrates two distinct patterns of allele dynamics over time. In the left column (infections p1, p66, and p140), we observe transient allele groups that emerge at one time point, with some reaching high frequencies before vanishing in subsequent time points (dashed lines). Consensus sequence samples from p1, p66, and p140 (as well as the other 6 infections shown here) form a monophyletic clade on a representative phylogeny of non-persistently infected individuals ^8^. Additionally, certain alleles that were not present at the early stages of infection surge to high frequencies towards the end of infection (bold solid lines). Conversely, the middle column (infections p132, p288, and p341) showcases alleles that experience a sweep from low to high frequencies, with some ultimately disappearing (dashed lines) and others reaching fixation (bold solid lines). The right column (p75, p288, and p523) show allele frequency dynamics that is a mix of the two patterns with some alleles appearing and disappearing in groups while other are present in the population in at least two time points, with some reaching fixation without disappearing at later time points.

**Supplementary Figure 4:**
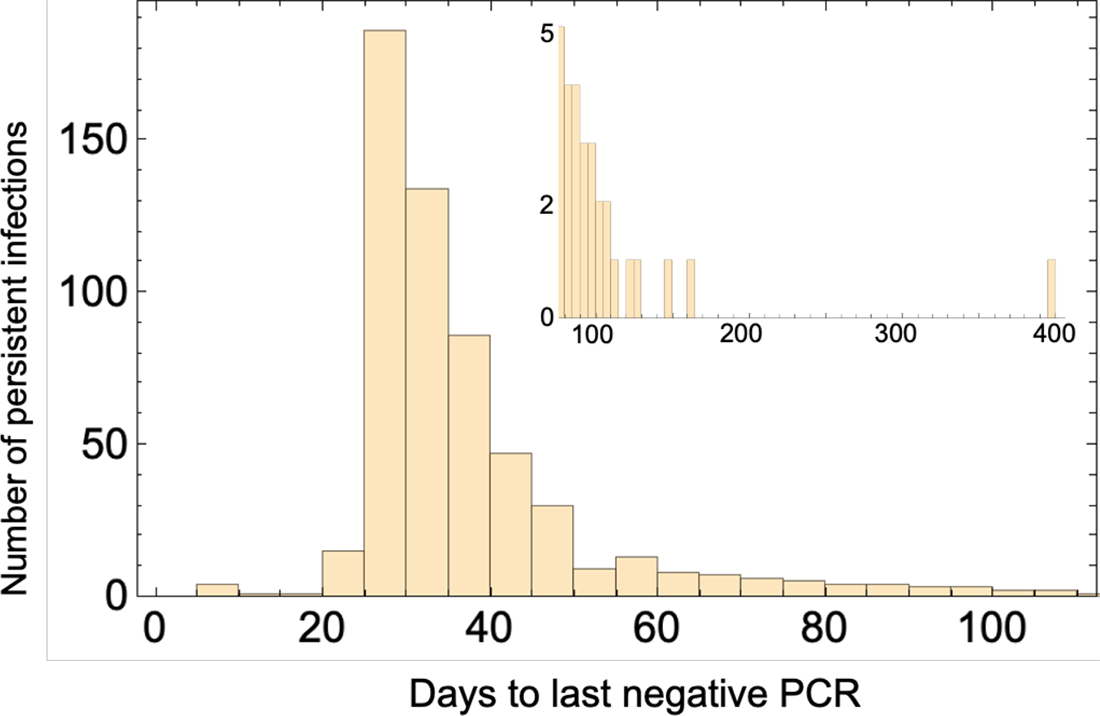
Number of days elapsed since the last time a persistently infected individual had a negative PCR test. The histogram plot includes all 576 identified persistent infections.

**Supplementary Figure 5:**
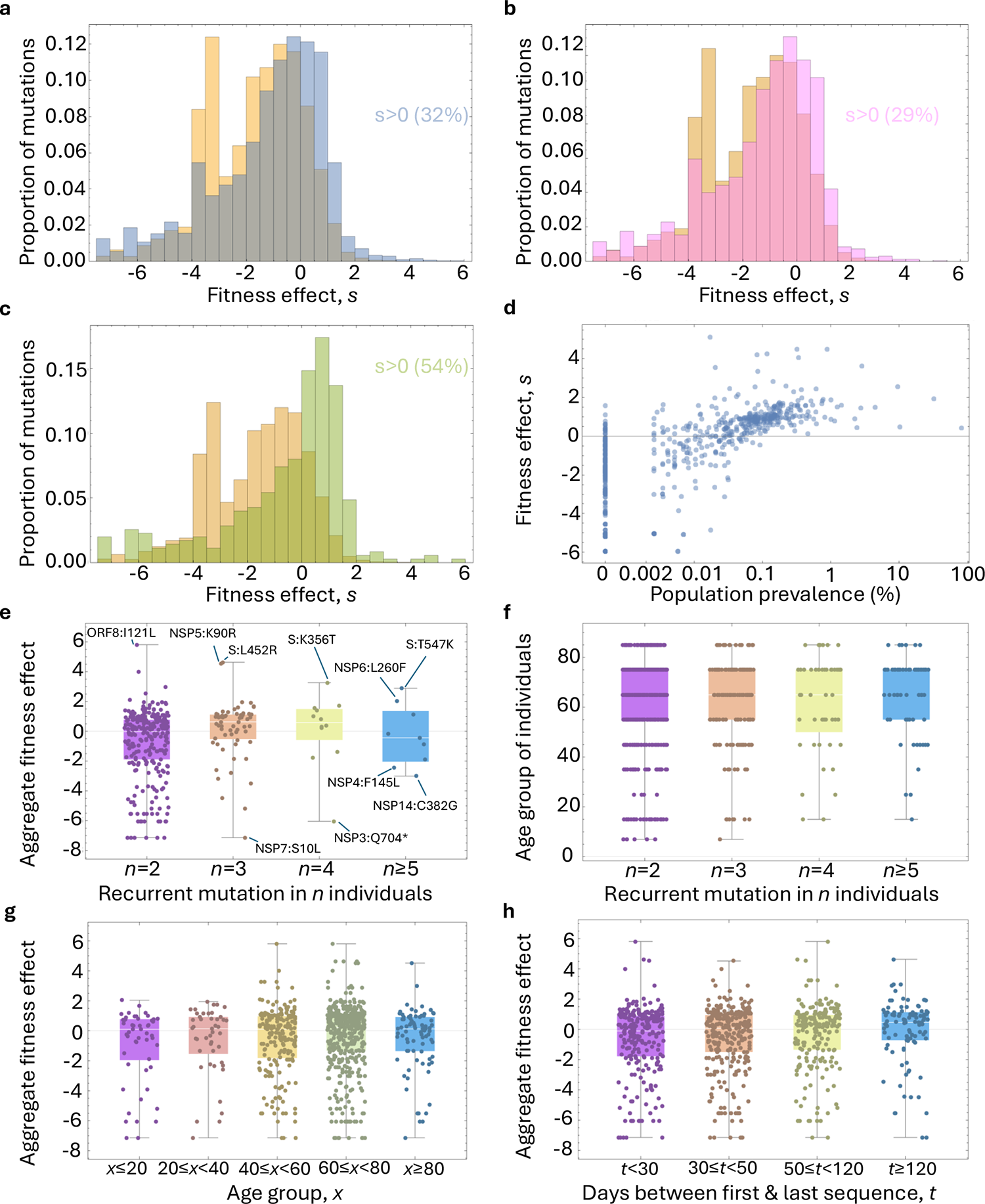
Between-host fitness effect and prevalence of recurrent mutations identified in persistently-infected individuals. **(a)** Distribution of between-host fitness effects of all SARS-CoV-2 mutations on a global phylogeny (orange), between-host fitness of all mutations found in persistently infected individuals (blue), **(b)** for those found only in a single persistent infection (magenta), and **(c)** for those found in two or more persistent infections (green). The percentage of mutations in persistent infections with a positive between-host fitness effect (*s*) is highlighted on each graph in (a)-(c). The between-host fitness effect of mutations in persistent infections corresponds to the fitness effect of that mutation on a global phylogeny within the same major viral lineage that was found to be in the persistently infected individual. For example, if a recurrent mutation is found in two persistently infected individuals with BA.2 and BA.5 infections, the between-host fitness effect of that mutation in both the BA.2 and BA.5 major lineages is recorded. **(d)** The between-host fitness effect of recurrent mutations found in persistent infections and their corresponding prevalence across all ONS-CIS sequences of the same major lineage as the persistent infection. **(e)** The aggregate between-host fitness effect (averaged across all major lineages of SARS-CoV-2 on a global phylogeny) of recurrent mutations found in *n* persistent infections. Some of the mutations with extremely high and low fitness effects are highlighted. **(f)** Age-group of all individuals which share *n* recurrent mutations. **(g)** Aggregate fitness effect of recurrent mutations per age group. **(h)** Aggregate fitness effect of recurrent mutations based on the duration of the persistent infection (as measured based on number of days between first and last sequence from a persistent infection) in which they emerged. Fitness effect of mutations are taken from https://github.com/jbloomlab/SARS2-mut-fitness/blob/main/results_public_2024-04-19/nt_fitness/ntmut_fitness_by_clade.csv ^25^.

**Supplementary Figure 6:**
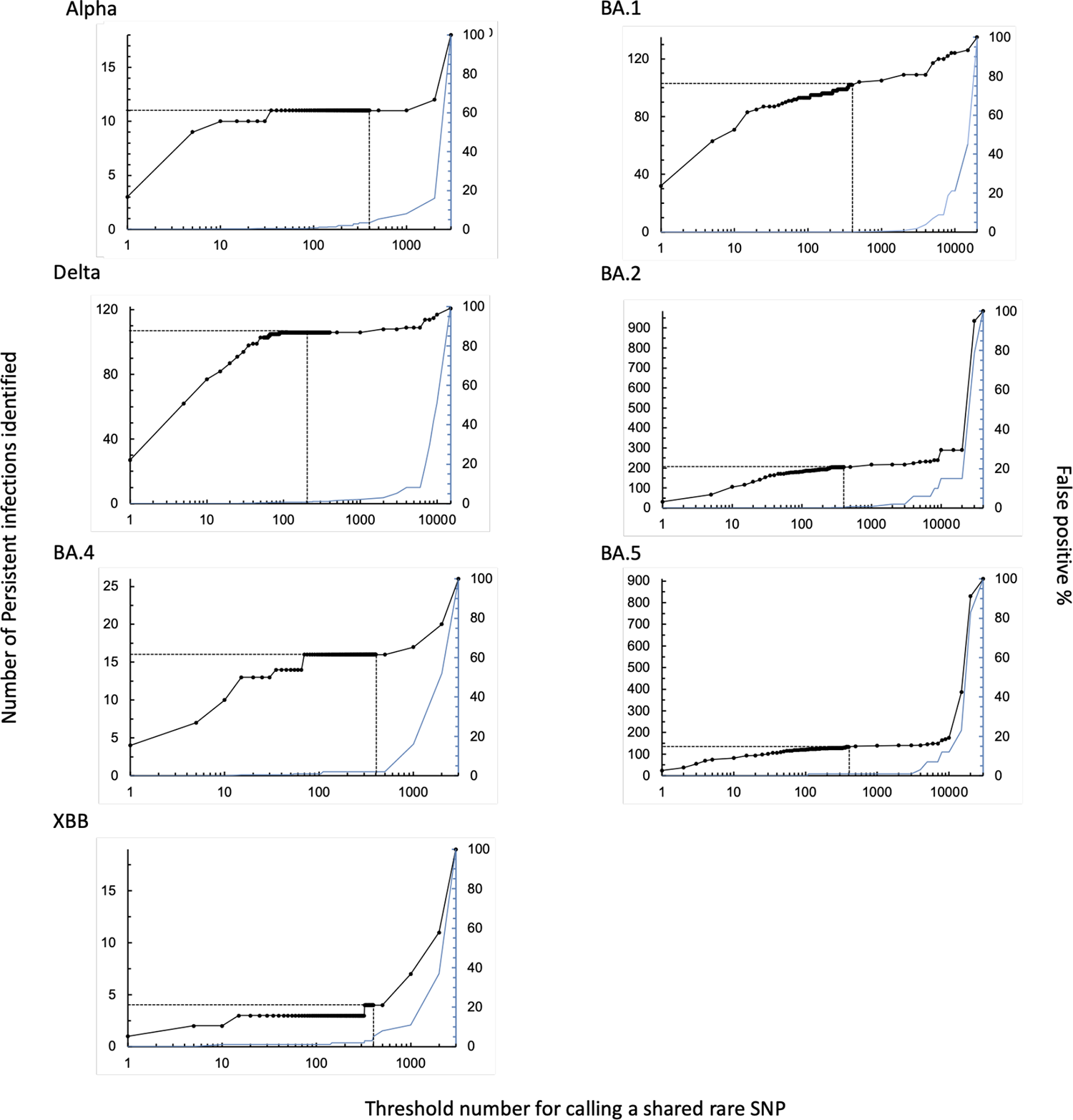
Number of persistent infections identified with a shared rare SNP as a function of the threshold number of cases for calling a rare SNP. A threshold value of 1 for a rare SNP means the rare SNP is only found in one sequence of that lineage in the ONS-CIS dataset, excluding sequences from any persistently infected individuals. The number of persistent infections identified gives the number of persistent infections lasting at least 26 days we would identify as persistent in the ONS-CIS using the given threshold (black). The false positive percentage gives the percentage of times two random samples of the same major lineage taken from the ONS-CIS would be falsely identified as belonging to the same persistent infection (blue; 1,000 pairs of samples were considered). As the threshold value for calling a rare SNP increases, the number of persistent infections identified (black) increases, but so does the false positive rate. Similar to the approach we took in our previous study ^8^, we chose a threshold number of 400 (vertical dashed line) in this study for identifying persistent infections, since for this threshold the percentage of false positives were 0-3% for all major lineages, but the number of persistent infections identified has begun to plateau. We allowed for possible misclassification of some BA.2 and BA.5 major lineages by allowing for potential identification of persistent infections with a mix of BA.2 and BA.5 samples.

**Supplementary Figure 7:**
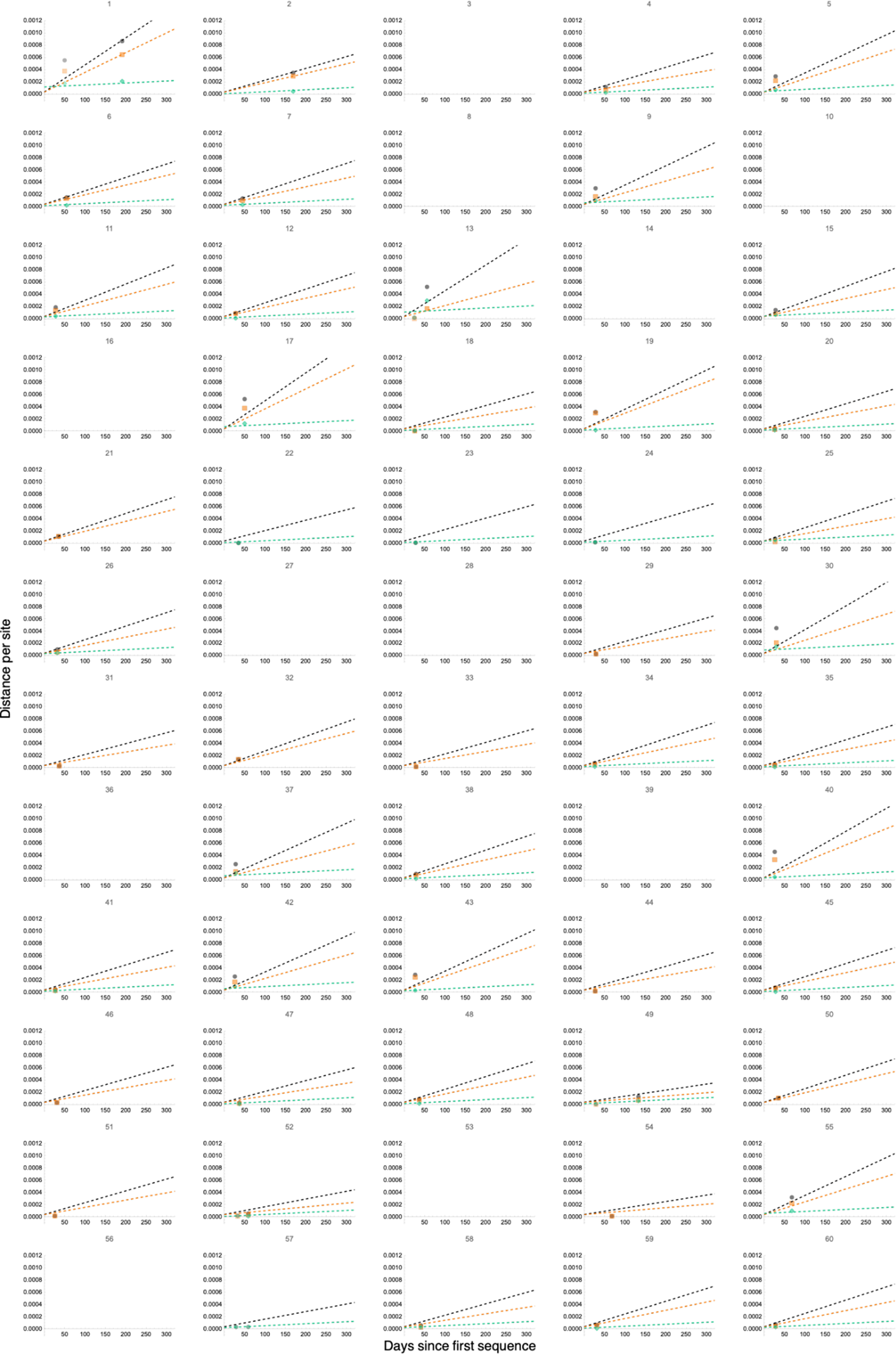
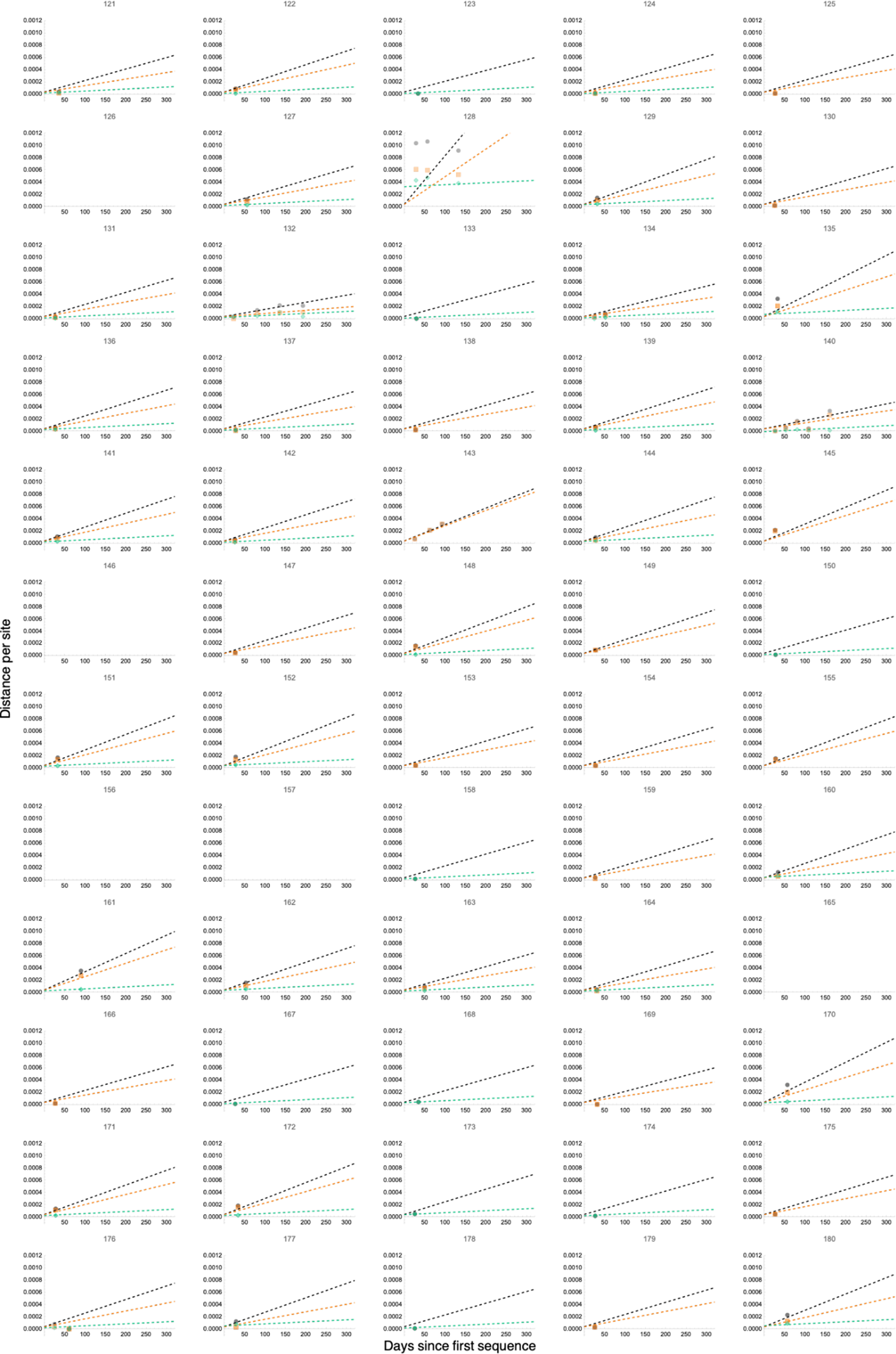
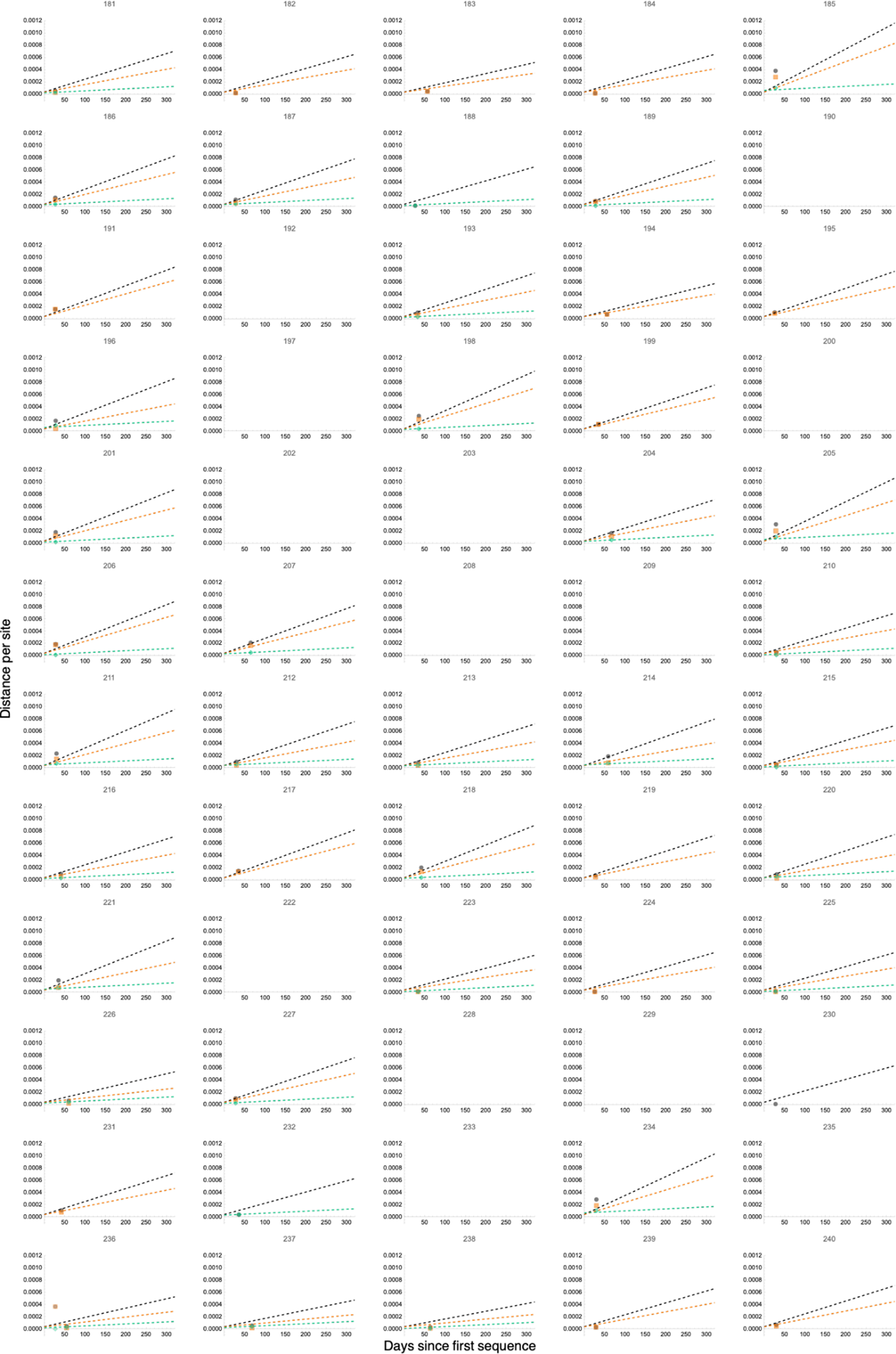
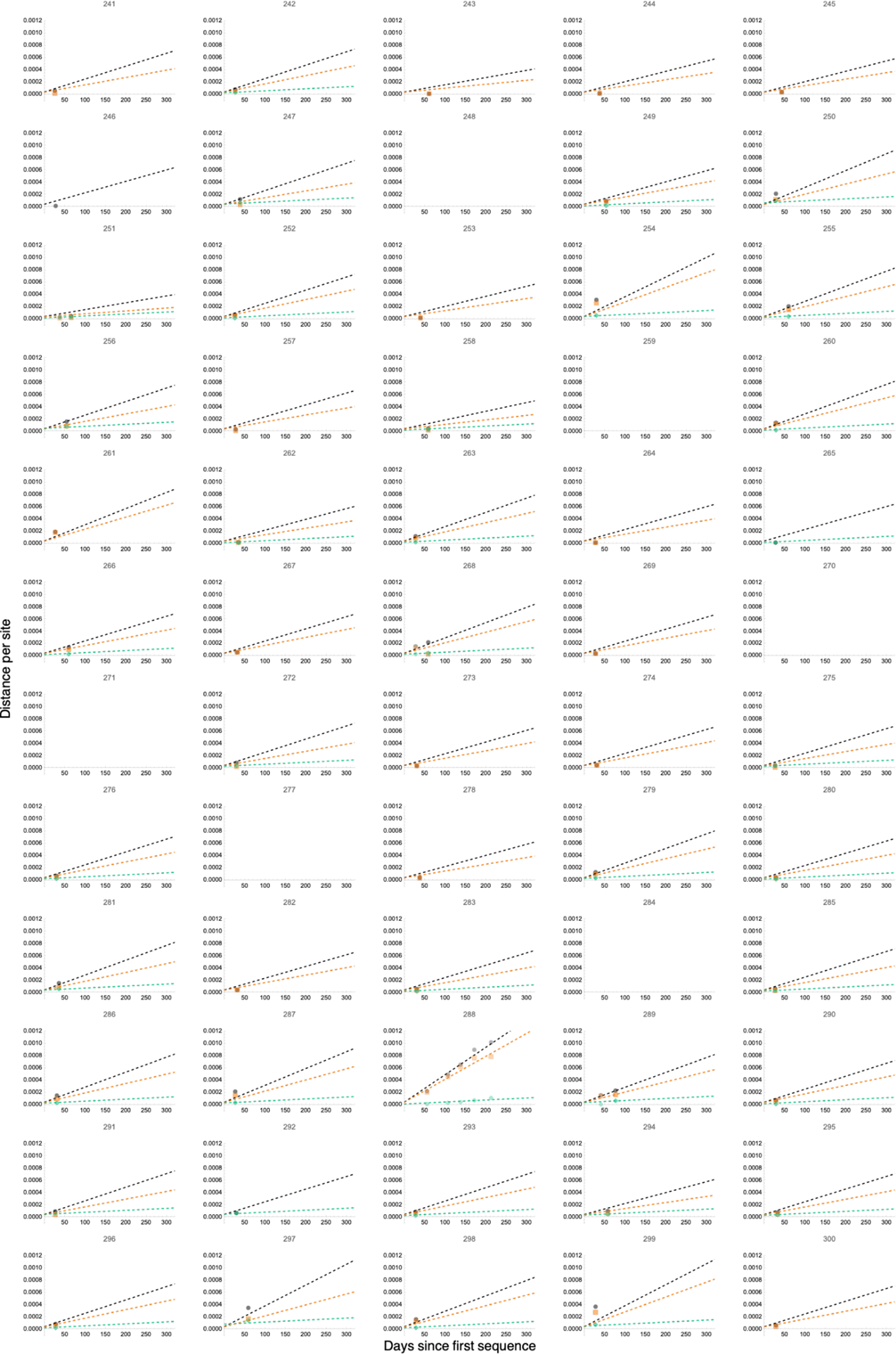
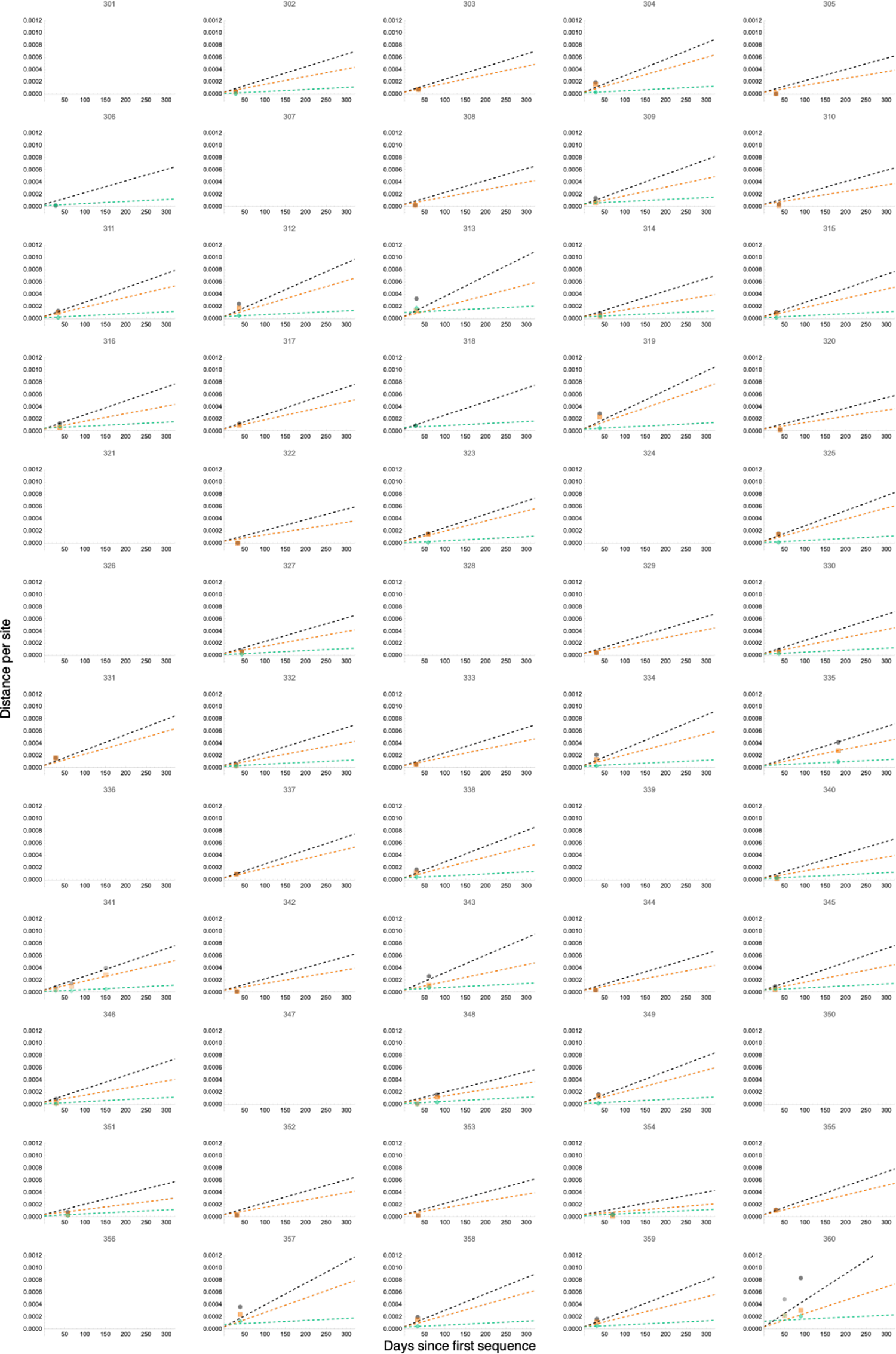
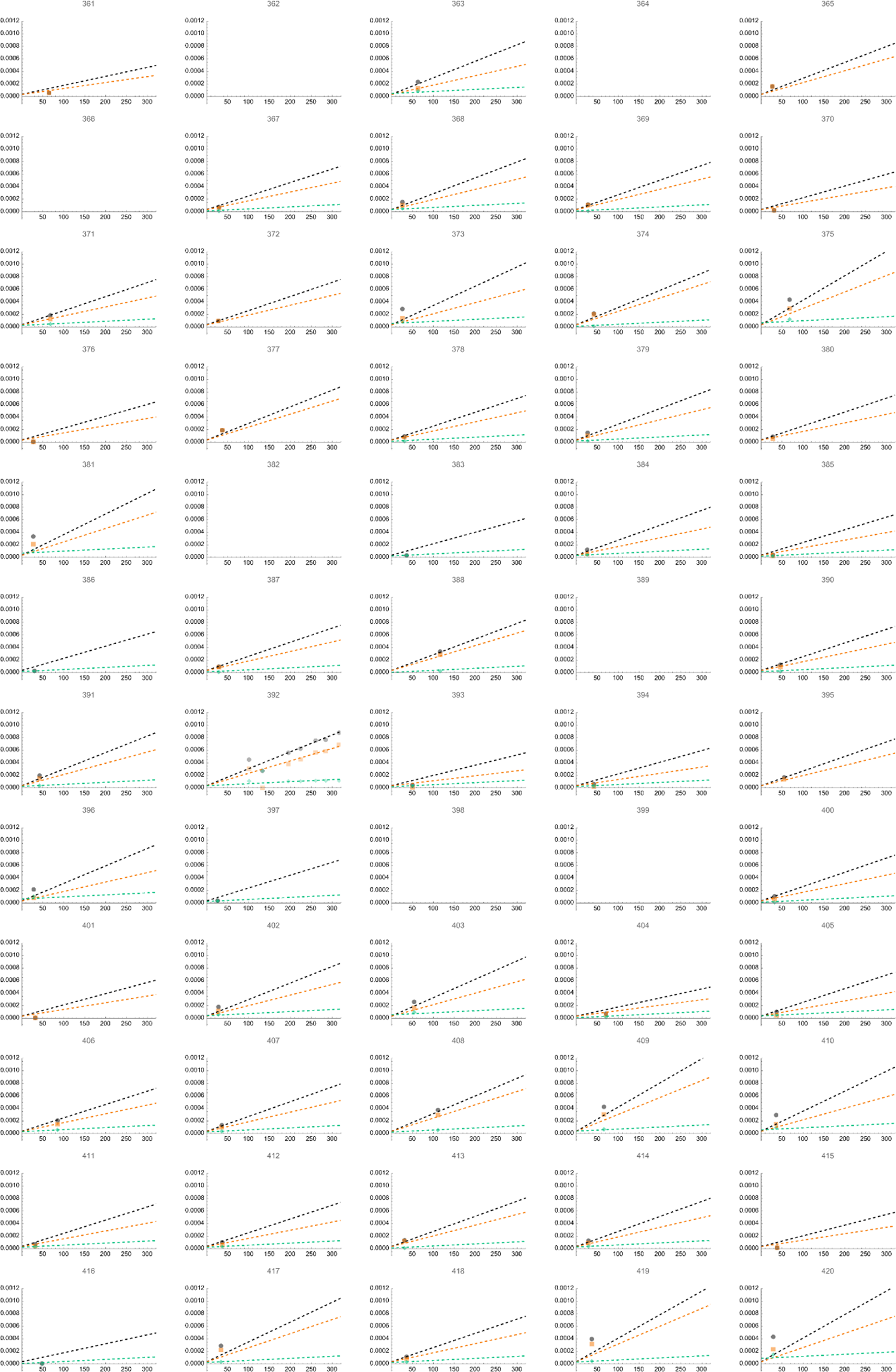
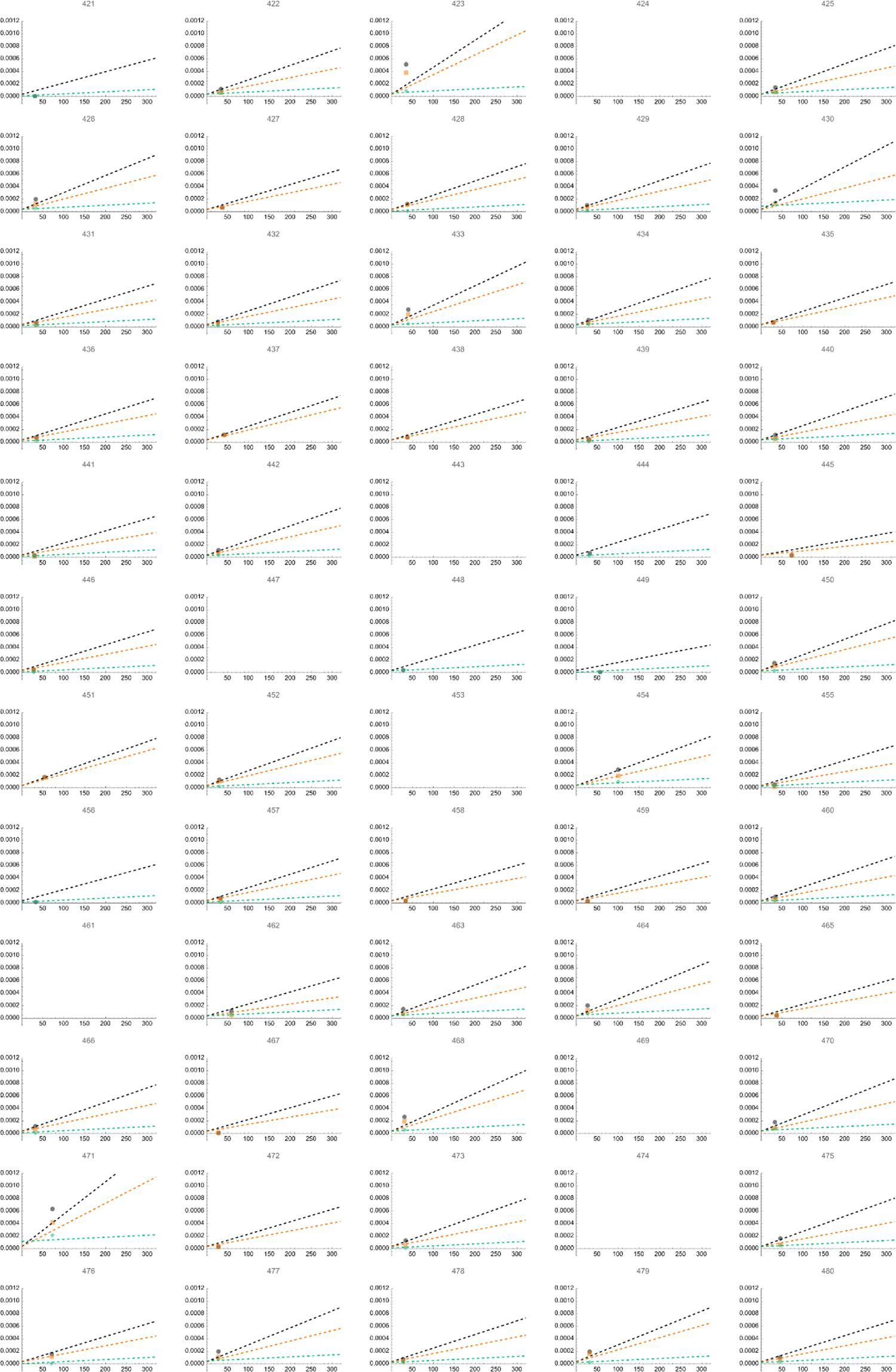
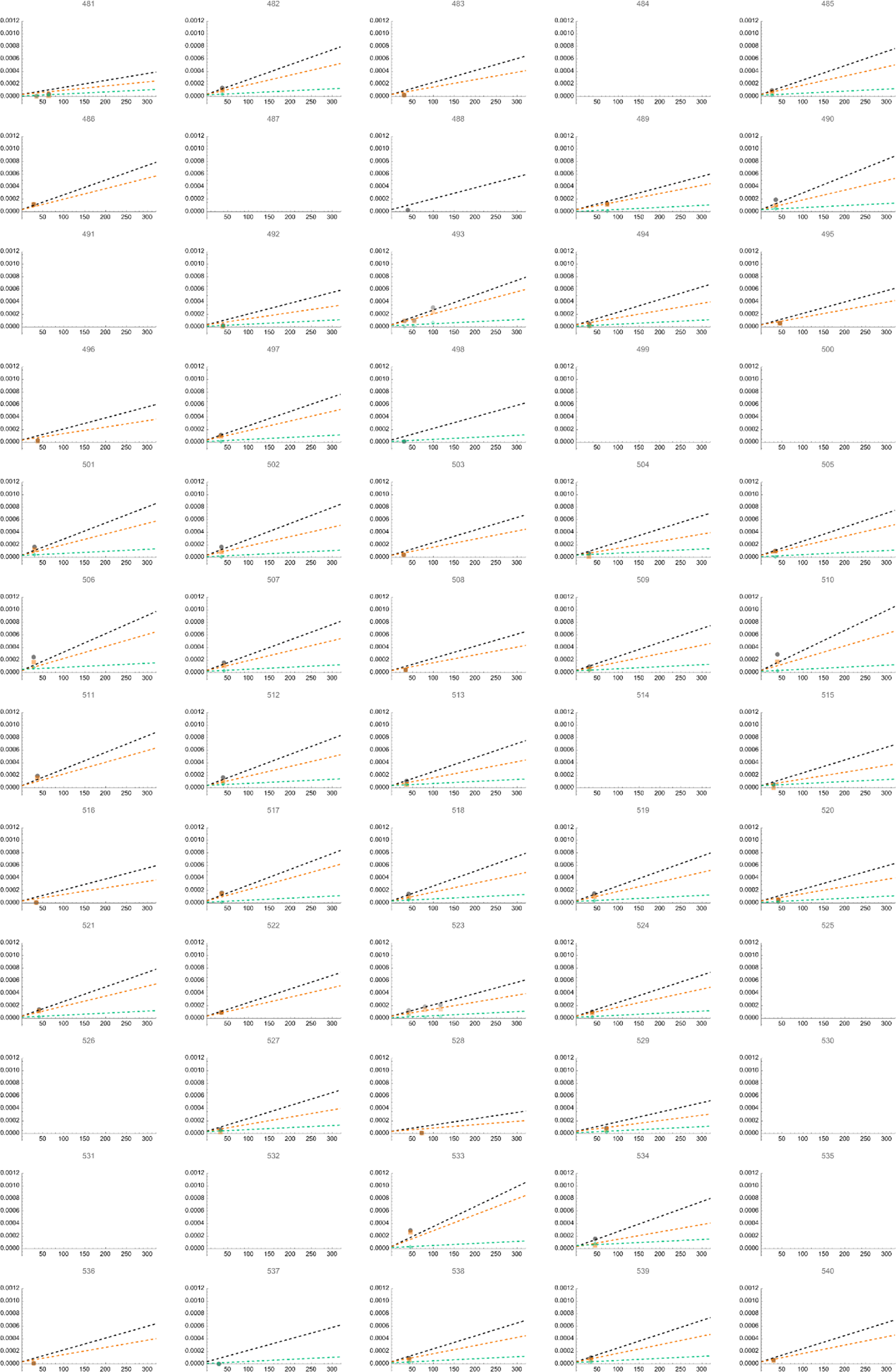
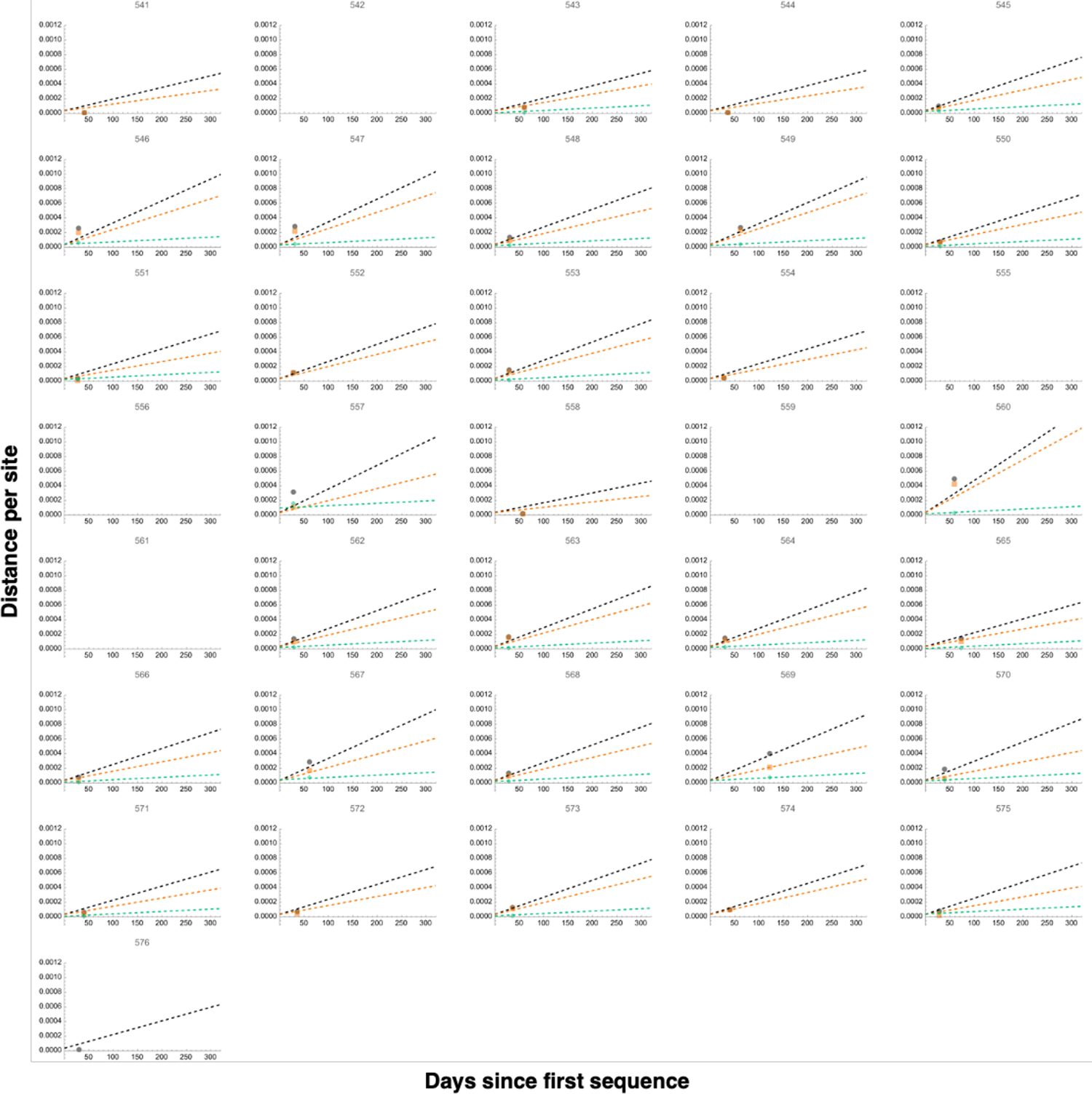
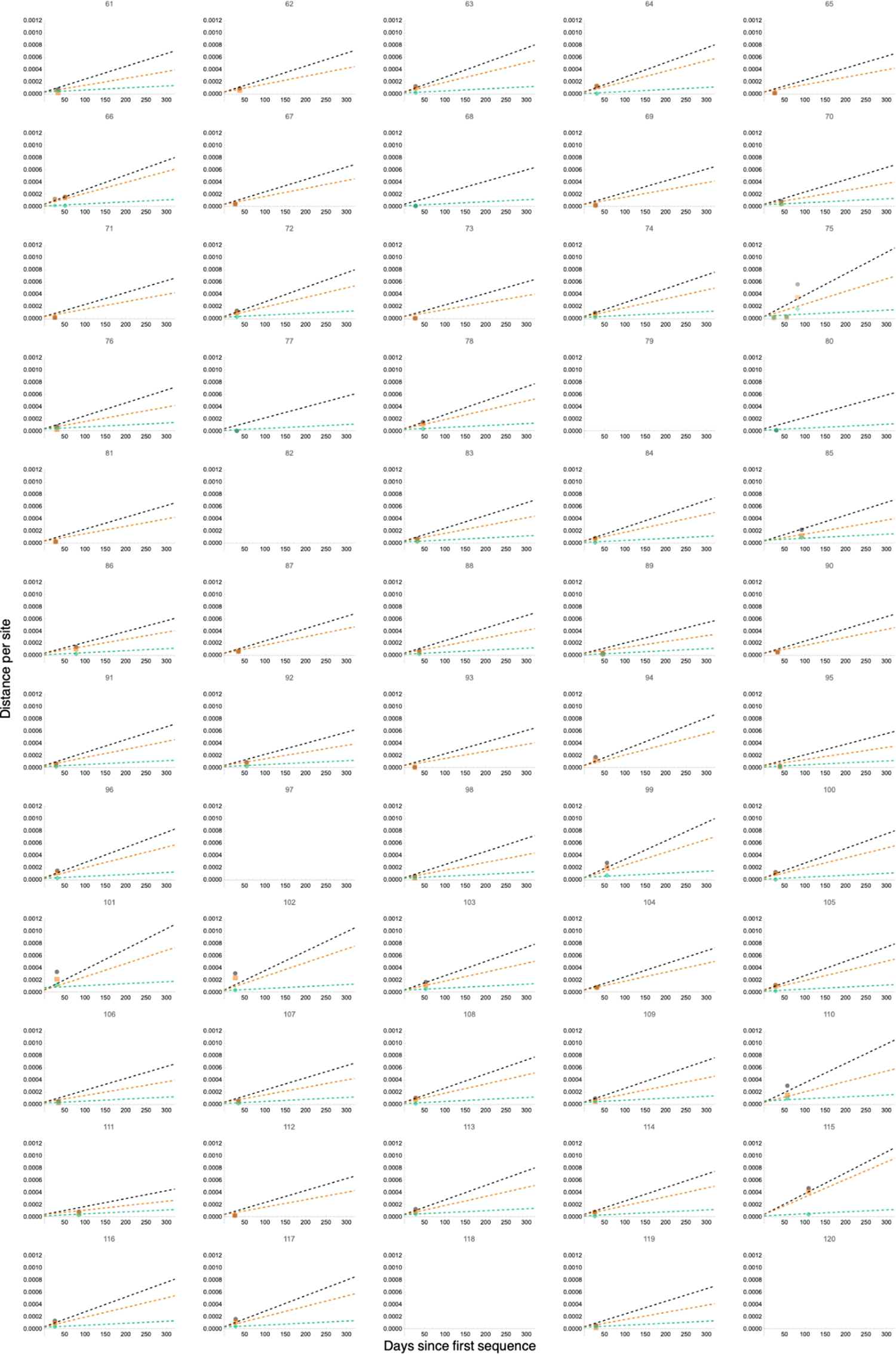
Rates of genome-wide, nonsynonymous, and synonymous evolution in all persistently infected individuals with measurable rates. The evolutionary distance over time for 494 persistently infected individuals with measurable genome-wide rate (black), 457 nonsynonymous rate (orange), and 368 synonymous rate (green). Points on the graph represent the total genetic distance from the consensus sequence at the initial time point, calculated based on allele frequency changes over time. Dashed lines indicate the regression lines that best fit these data.

